# High-resolution characterization of nasal microbial dynamics in young children

**DOI:** 10.1101/2025.08.22.25334231

**Authors:** Dennis Hoving, Rosanne A.M. Steenbergen, Youvika Singh, Dima El Safadi, Ester L. German, Ivana Barros de Campos, Elandri Fourie, Mara van Roermund, Alicia C. de Kroon, Iris van der Valk, Debby Bogaert, Els Wessels, Jutte J.C. de Vries, Willem R. Miellet, Krzysztof Trzciński, Daniela M. Ferreira, Marlies A. van Houten, Simon P. Jochems

## Abstract

Nasal carriage of bacteria and viruses is common in young children. However, these carriage dynamics remain poorly understood due to the lack of high-resolution studies. Therefore, we conducted a 29-day prospective study in which 45 children, 1-5 years of age, provided daily nasosorption samples from which we analyzed 16 viruses, 6 bacteria, and 21 *Streptococcus pneumoniae* serotypes/groups. Bacterial densities fluctuated rapidly, with autocorrelations lasting only eight days. Inter-species but also intra-species interactions were observed; co-carriage of multiple *Streptococcus pneumoniae* serotypes increased clearance of non-dominant strains. *Staphylococcus aureus* carriage was associated with reduced viral acquisition and accelerated viral clearance. Rhinovirus triggered rhinitis on the day of acquisition, with bacterial densities rising three days later. This high-resolution sampling approach elucidated complex microbial and host-pathogen interactions in children.

## Introduction

Respiratory tract infections are a global health problem, causing more than two million deaths each year, especially in infants and older adults (*1*). Young children are frequently colonized by, or carry, bacteria that can cause disease, like *Streptococcus pneumoniae, Staphylococcus aureus, Haemophilus influenzae* and *Moraxella catarrhalis* (*2*). Moreover, viruses are frequently detected from nasal samples, even in children without overt signs of infection (*3*), a phenomenon called viral carriage. Bacterial and viral carriage episodes are a driving factor for community transmission (*4*), can lead to rhinitis (*5*), and are also a prerequisite for invasive disease for many of these pathogens (*6*).

During carriage, bacteria and viruses show associations with each other and the host, as demonstrated by epidemiological studies (*3*). For example, viral carriage is often linked to increased *S. pneumoniae, H. influenzae* and *M. catarrhalis* density, while *S. aureus* is negatively associated with these three bacteria. To understand these interactions, many pre-clinical studies in animal models and controlled infections studies in human adults have been performed, identifying direct interactions between microbes but also interactions mediated by altered host responses (*7–10*). However, the extent to which such findings from adults or animal studies translate to children remains unclear.

Collecting longitudinal data at high temporal resolution in children could elucidate the directionality of host-pathogen and microbe-microbe interactions. The main obstacle to collecting high-frequency data from children has been that classical sampling methods, such as nasopharyngeal swabs, are typically not well tolerated (*11*). Recently, alternative less-invasive sampling methods have been developed, such as the nasosorption sample, a synthetic absorptive matrix that is placed in the nose and absorbs epithelial lining fluid (*12*). Previous studies have shown that such nasosorption samples can be used to study microbes in the nose of children. They show an excellent correlation with nasopharyngeal swabs for *S. pneumoniae* carriage detection, a similar total microbiota composition compared to nasal washes in healthy children (*13, 14*), and detect RSV comparably to nasal aspirates in infants a paediatric ICU setting (*15*).

To provide a better understanding of microbial dynamics and host-microbial interactions, we conducted a prospective cohort study, including 45 young children with daily nasal sampling performed by parents for 29 days and analysed viral and bacterial dynamics. We aimed to elucidate directionality of microbial associations and link these to symptom development using this high-frequency sampling approach. Overall, this study provides multiple new insights and opens up new avenues of investigating host-pathogen interactions at the nasal mucosa in children.

## Results

### Cohort characteristics and daily sampling

Between January 2022 and October 2023 we enrolled 46 healthy children, of which 45 completed the 29-day study (97.8%) (**Fig. S1**). These 45 children included 28 girls and 17 boys, were aged 1-5 years old, and all attended school or day care at least two days a week (**Fig. 1A**). Parents collected daily nasosorption samples from their children and filled out questionnaires about sample collection, symptoms, activities and medication usage. In total 1262 nasosorption samples were collected (average 28.0/child, range 22-29/child), while the questionnaire was fully completed 1184 times (average 26.3/child), indicating good adherence to the study protocol.

**Figure 1.**
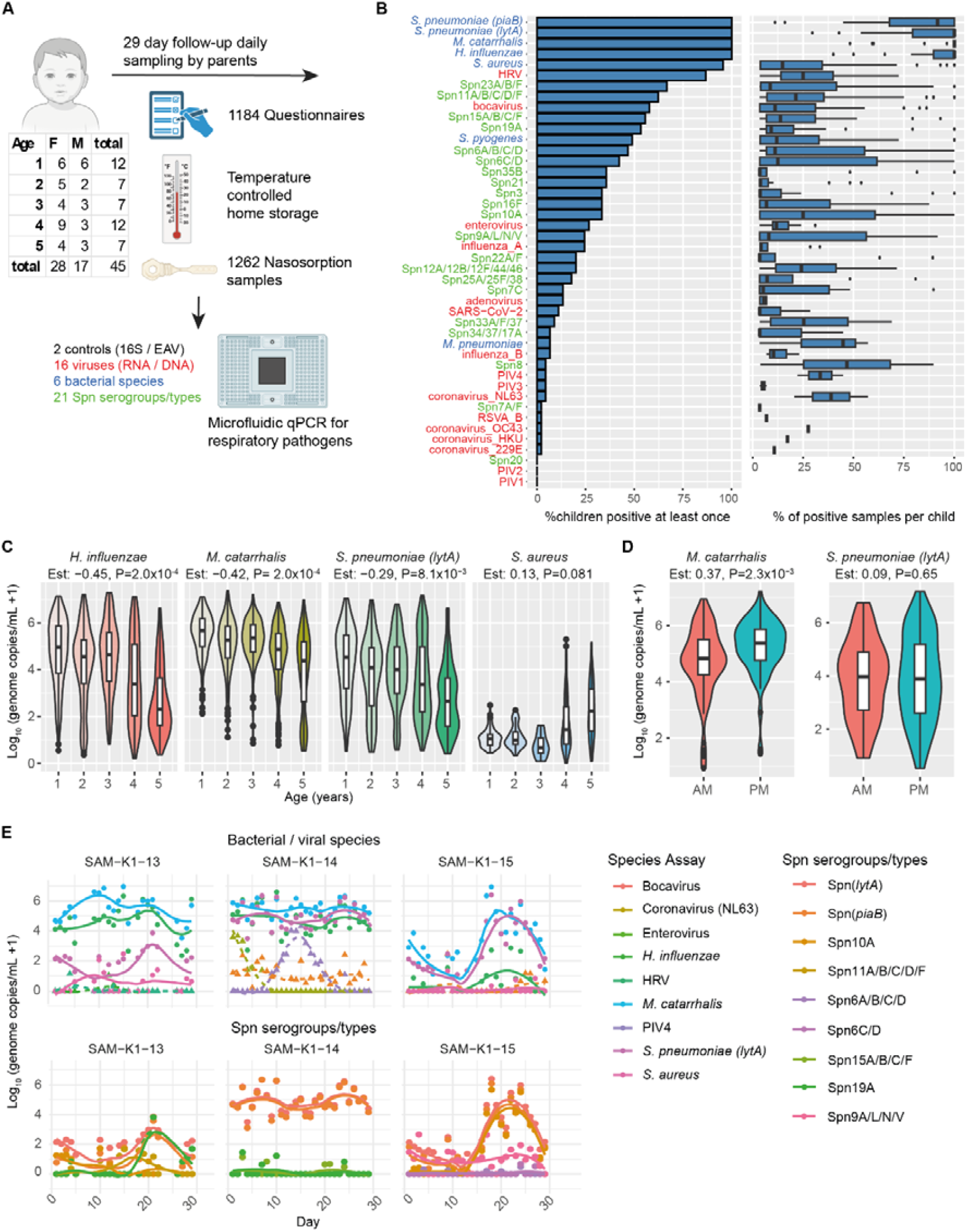
Cohort and microbial detection. **A)**. Schematic overview of samples and questionnaires collected, microbial analysis and table showing age and sex of included children. **B)** Bar plot and box plot showing the percentage of children (left) and samples per child (right) that are detected per assay. Assay names are colored by type of pathogen: bacterial species (blue), *S. pneumoniae* (Spn) serotypes (green), viral (red) **C)** Pathogen density for most common bacterial targets stratified by age. Statistical results from linear mixed model including child ID as random effect and age as linear fixed effect estimate (Est) are shown. Only samples with detectable bacterial loads are included. **D)** Log-transformed density of *M. catarrhalis* and *S. pneumoniae* in samples collected in morning (AM) or afternoon/evening (PM) are shown. Only samples with detectable bacterial density and samples on day before, day of or day after change in collection period are included. Statistical results from linear mixed model including child ID as random effect and collection period as fixed effect (Est) are shown. **E)** Longitudinal analysis of bacterial (continuous lines and circles) and viral (dashed lines and triangles) carriage densities (top) or Spn serotypes (bottom) are shown for 3 representative children. Individual points and loess curves are shown. Per child only assays that were carried at any point are shown, with carriage defined as having at least two positive samples in a row allowing for a gap of three negative samples in between.

To assess tolerability of the repeated daily sampling, we evaluated an exit questionnaire after the first 10 included children and after the remainder of the cohort (**Fig. S2**). In the pilot cohort 2/10 parents (20%) found the study to be taxing, therefore we reduced nasosorption sampling time from 60 seconds to 30 seconds, in line with previous pediatric studies (*15, 16*). This did not affect bacterial or viral detection (**Fig. S3A**,**B**). After reducing the sampling duration, only one of the 29 parents who filled out the questionnaire found it taxing (3.4%, **Fig. S2**). Samples collected by parents showed similar microbial densities compared to samples collected by clinicians, demonstrating successful self-sampling (**Fig. S3C**).

We used a microfluidic qPCR to simultaneously quantify 16 viruses, 6 bacterial species and 21 *S. pneumoniae* serotypes/groups (**Table S1 and Fig. S4**). We also included a RNA virus spike-in technical control and included 16S for overall bacteria as a sample collection control. Of all microbial assays, only 3 were never detected: parainfluenza virus (PIV)1, PIV2 and *S. pneumoniae* serotype 20 (**Fig. 1B**). *S. pneumoniae* serotype detection strongly correlated with the distribution of concurrently circulating serotypes in the Netherlands (**Fig. S4B**)(*17*). As expected, given the inclusion of only daycare/school-going children, the bacteria *M. catarrhalis, S. pneumoniae* and *H. influenzae* were found in samples from all children at some point during the study, and were detected in the majority of samples per child (*18*). *S. aureus* was detected in samples from 95.6% of children, but the number of positive samples varied greatly per child. Some children showed *S. aureus* positivity throughout the entirety of the sampling period, while the majority of children had *S. aureus* detection in less than 1/3 of the collected samples, in line with literature that *S. aureus* is often carried intermittently (*19*). *S. pyogenes* and *M. pneumoniae* were found in samples from 48.9% and 6.7% of children, respectively. Of the viral pathogens, human rhinovirus (HRV) was detected in samples from 86.7% of children, bocavirus in 57.8%, enterovirus in 26.7%, influenza A in 24.4% and all other viruses in 13% or less.

Bacterial density decreased with age for *M. catarrhalis, S. pneumoniae* and *H. influenzae*, but not for *S. aureus*, based on a mixed effect model to account for the repeated sampling per child (**Fig. 1C**). This also agrees with existing literature that shows comparable age-associated variations in bacterial density (*20*). Although parents were instructed to collect samples at the same time of day as much as possible, samples were collected in morning (16.6%, n=204), afternoon (13.0%, n=160) and evening (70.3%, n=864). We observed that for *M. catarrhalis*, but not for any of the other bacteria, density fluctuated based on collection period (**Fig. S5**). Densities were lower in samples collected during the morning compared to afternoon or evening. This effect remained when restricting our analysis to days surrounding changes in sample collection period to reduce noise (estimate=2.3x lower in morning, p=0.002, **Fig. 1D**).

Thus, daily, minimally-invasive sampling in young children by parents was feasible, and allowed for the observation of previously known and unknown features of microbial carriage.

### Daily sampling reveals microbial dynamics and exposure events

Longitudinal sampling allowed us to track bacterial and viral carriage over time and observe their dynamics, including acquisition and clearance events (**Fig. 1E, Fig. S6** and **Fig. S7**). Moreover, through daily sampling we observed exposure events (**Fig. 2A**). As expected, exposure events associated with same-day school or daycare attendance (adjusted odds ratio, OR, 1.60, p=3.6×10^−5^). They also associated with other reported social activities involving other children, such as birthday parties (adjusted OR 1.68, p=0.003), although they did not associate with having gone to the swimming pool (adjusted OR 0.92, p=0.75, **Fig. 2B**). The association of exposure with known places where transmission occurs, provides confidence that we were detecting true new exposures in children. In total, we identified 451 exposure events in the entire cohort (average 10.0/child during the follow-up). Of these exposure events, 241 (53.4%) occurred in isolation, as a single new microbe was observed. In all other cases, from two up to five microbes were simultaneously newly detected (**Fig. 2C**). During the 29-day study period, a child was exposed on average more than once to any *S. pneumoniae* serotype (5.4 times), *S. aureus* (1.2 times) and any virus (2.4 times, **Fig. 2D**). For all viruses combined, exposure rates decreased with age, which was partly explained by decreasing exposure to bocavirus (**Fig. 2E**).

**Figure 2.**
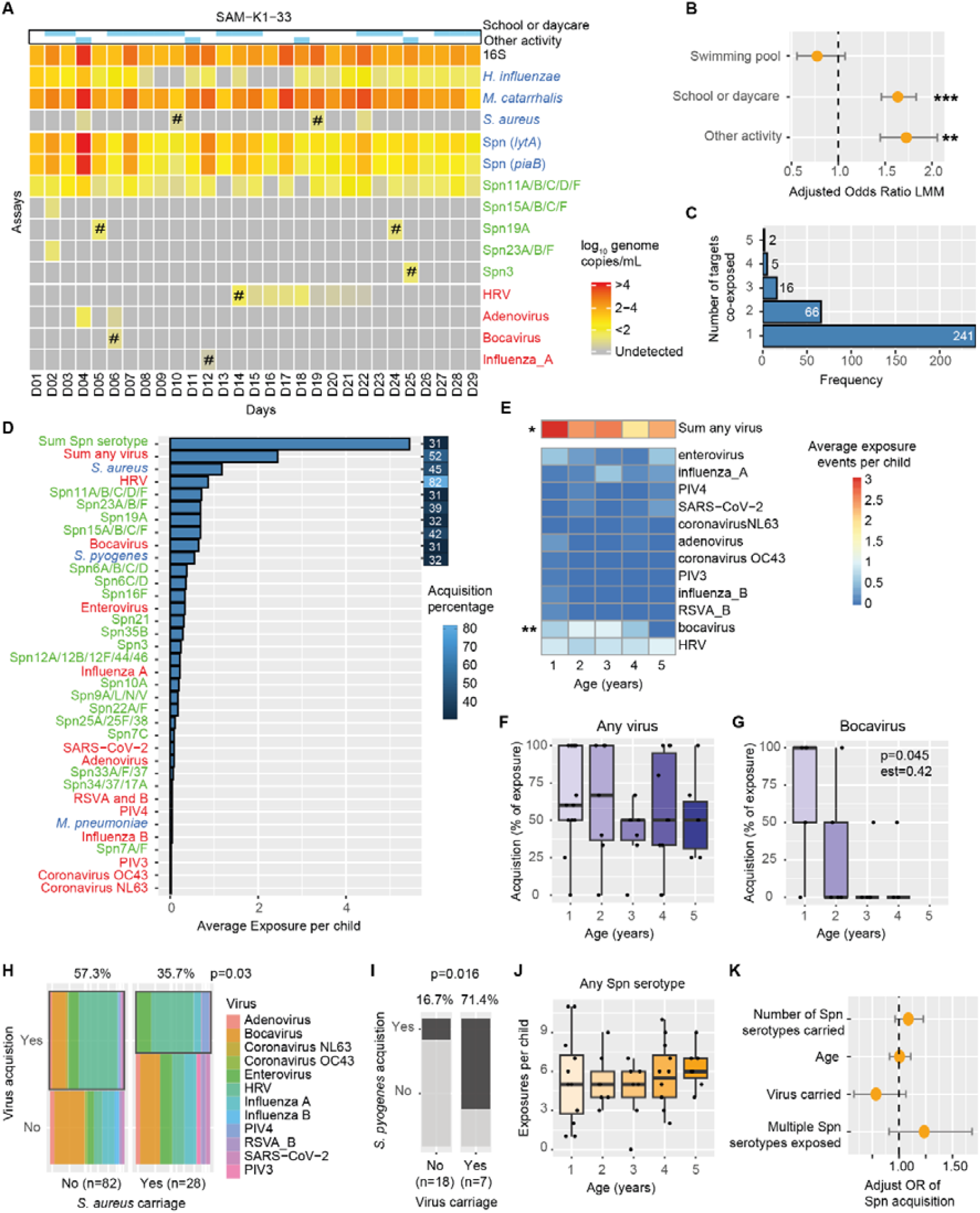
Exposure and pathogen acquisition in children. **A)**. Heatmap showing for one child the densities of all qPCR targets for which there was at least one positive sample (rows) per day (columns). Grey indicates undetectable. Top 2 rows indicate the filled in questionnaire questions about potential exposure events. # represents an exposure event defined as having a positive qPCR assay result after having at least 4 consecutive negative samples. **B)** Association between social activities and detection of exposure events. Only samples collected in afternoon or evening were included to prevent associating newly detected microbes in the morning with activities occurring later in time. Estimate and standard errors are shown from a binomial linear mixed model including assay and child as random variables and the three social activities as fixed effects. **P<0.01, ***P<0.001. **C)** Bar plots indicating how often exposure of multiple targets at the same timepoint occurred. **D)** Bar plot indicating how often on average a child showed exposure for an assay during the 29 day follow-up. The sum of all *S. pneumoniae* (Spn) serotypes and the sum of all viral targets are included. The assays for *Haemophilus influenzae, Moraxella catarrhalis*, Spn *lytA* and *piaB* (overall Spn) are excluded as they were so common that exposure could not be defined. The heatmap next to the barplot indicates the percentage of time in which an exposure resulted in carriage, defined as having at least two positive assays with no more than 3 negative days in between. Acquisition percentages are only shown for targets with at least 20 exposure events. **E)** Heatmap showing exposure rates for all viral targets, stratified by age. **P<0.01 from binomial linear mixed model including child ID as random effect and age as linear fixed effect. Only all viruses, bocavirus and rhinovirus were tested. **F, G)** Boxplots showing the success rate of acquisition after exposure for all viruses and bocavirus only, stratified by age. Individual children are depicted as dots. Estimate and p-value of binomial linear mixed model including only exposure events with acquisition success as outcome variable and age as linear fixed effect and child ID as random effect. **H)** Mosaic plot indicating the acquisition percentage of viral exposures in children stratified by concurrent *S. aureus* carriage or not. Colour indicate the encountered viral pathogens and the percentage of successful acquisition is shown. P-value from binomial mixed effects model, including *S. aureus* carriage as fixed effect and viral assay and child ID as random effects. **I)** Acquisition success of *S. pyogenes* of children with concurrent viral infection or not. Percentages of acquisition and p-value of binomial mixed effects model are depicted. **J)** Exposure events of Spn serotypes, stratified by age. Boxplots and individual children are depicted. **K)** Point estimates and standard errors of binomial mixed effect model assessing the effect of the number of pre-existing Spn serotypes, age, viral carriage and the exposure to more than 1 serotype simultaneously (fixed effects) on *Spn* carriage acquisition, including exposed serotype and child ID as random effects.

We then assessed whether exposure led to carriage, defined as a pathogen being detected at least twice in a five-day period (**Fig. 2D)**. For HRV, 82% of exposure events led to carriage acquisition, while for bocavirus this was only 31%. Exposure to bacteria *S. aureus* (45%), *S. pyogenes* (32%) and cumulative *S. pneumoniae* serotypes (31%) led to acquisition less frequently than for all viruses (57%). While the probability of carriage acquisition upon exposure to all viruses combined was not affected by age, susceptibility to bocavirus acquisition decreased with increasing age (p=0.045, **Fig. 2F,G**). Children who were carrying *S. aureus* at the time of viral exposure were less likely to become viral carriers than children without *S. aureus* (35.7% versus 57.3%, p=0.030, **Fig. 2H**). In turn, viral carriage also had an effect on bacterial acquisition: *S. pyogenes* acquisition was more likely in children carrying a virus at the day of exposure than in children without a virus (71.4% versus 16.7%, p=0.016, **Fig. 2I**). We also assessed whether exposure or carriage acquisition of *S. pneumoniae* could be explained by co-factors. Analysis was done at a serotype level as most children were positive for the general *S. pneumoniae* genes *lytA* and *piaB* in most samples. *S. pneumoniae* exposure did not change with age (**Fig. 2J**). Moreover, *S. pneumoniae* acquisition probability was not associated with age, viral carriage, being exposed to more than one serotype, or with the number of already carried *S. pneumoniae* serotypes (**Fig. 2K**).

Thus, through daily sampling we were able to define exposure events and susceptibility to carriage acquisition, which depending on the microbe encountered could be associated with age, or already carried other bacteria or viruses.

### Bacterial inter-species interactions and stability

To get better insight into bacterial dynamics and interactions we focused on the most commonly present bacteria: *S. aureus, M. catarrhalis, S. pneumoniae* and *H. influenzae*. One child received antibiotics (Phenethicillin) for a throat infection, which led to a transient decrease in *S. pneumoniae* density, although carriage was not completely cleared and density rebounded to >10^6^ copies/mL in just two days after the antibiotics course finished (**Fig. S8A**). Another child received antibiotics (Nitrofurantoin) for a urinary tract infection. To avoid potential effects on bacterial interactions, we removed all samples collected during antibiotics treatment from subsequent analyses. Performing a partial correlation analysis, which identifies correlations between parameters while controlling for the other included variables, indicated that *M. catarrhalis, S. pneumoniae* and *H. influenzae* were positively associated with each other, while *S. aureus* was negatively associated with *M. catarrhalis* (**Fig. 3A**), in line with other studies (*3*). To get a better insight into these interactions, we used a multi-level vector autoregression model (*21*), which separates partial correlations into within-subject effects (comparing fluctuations within an individual) and between-subject effects (comparing mean between individuals). This revealed that the negative association between *S. aureus* and *M. catarrhalis* was mostly due to between-subject effects, while no negative association between them was apparent within individuals (**Fig. 3B, C**). The positive association between *S. pneumoniae, M. catarrhalis* and *H. influenzae* was seen both within and between individuals (**Fig. 3B, C**). Repeating these analyses with Loess estimates or 16S-normalized densities rather than directly measured concentrations, in order to address potential sampling effects, confirmed these findings (**Fig. S8B-E**). To assess whether these bacterial interactions were driven by concurrent viral carriage, we also performed analysis in samples without concurrent viral carriage (**Fig. S8F**,**G**). In the absence of virus, the correlation structure between bacterial species was not affected, suggesting that direct interactions between bacteria are not completely mediated by viral effects.

**Figure 3.**
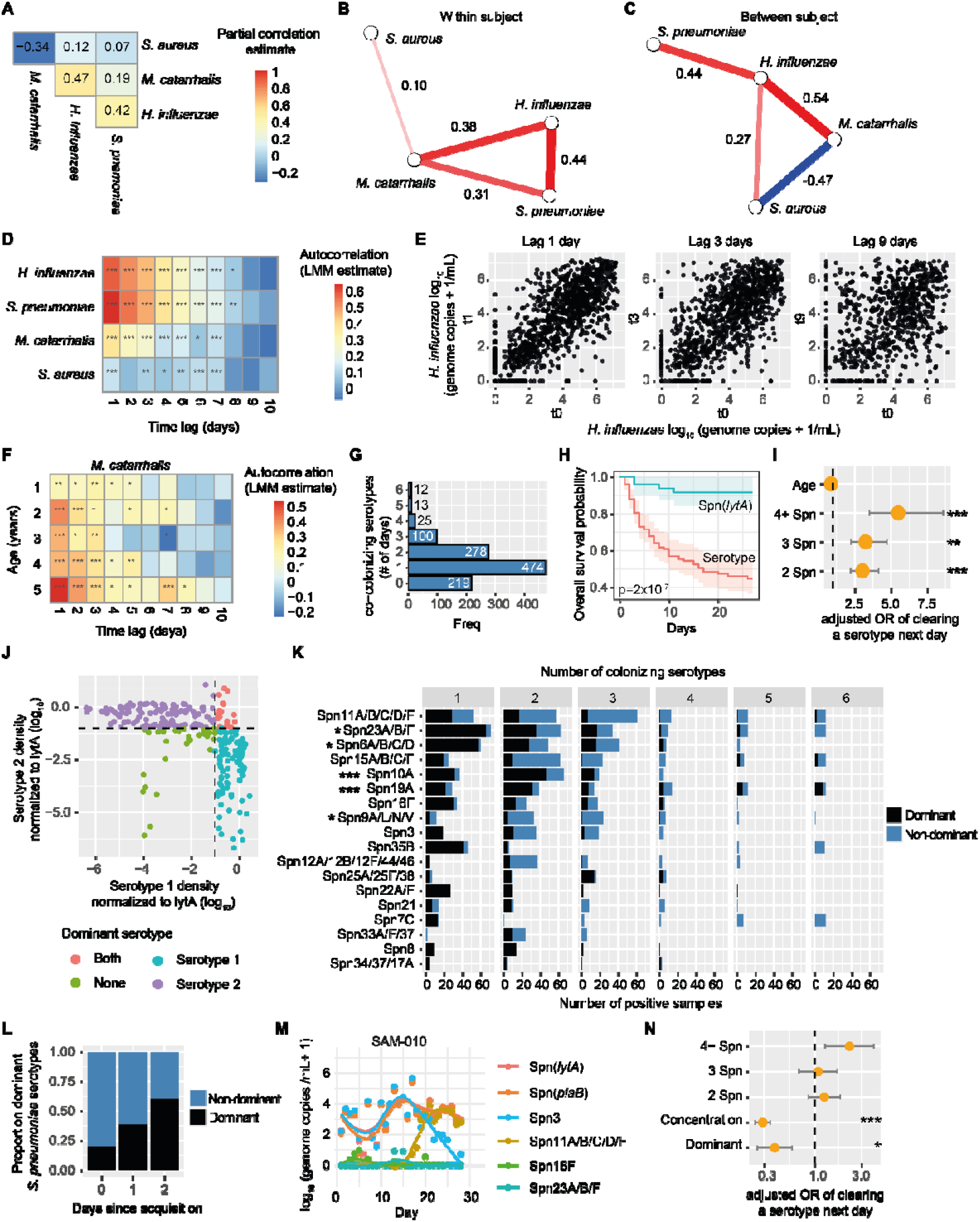
Bacterial interactions and carriage dynamics in children. **A)** Heatmap of partial correlations of log-transformed bacterial densities. **B)** Multi-level vector autoregression modelling showing within subject and **C)** between subject partial correlations between bacterial species. Red and blue lines indicate positive and negative associations respectively, with only lines with P<0.05 depicted. **D)** Autocorrelation for log-transformed densities per species, using a linear mixed effect model (LMM) including child as random variable. Columns indicate time lag. Correction for multiple testing was done using Benjamini-Hochberg correction. **E)** Example dot plots for the correlation between 1, 3 or 9 days lag in *H. influenzae* density. **F)** Heatmap showing autocorrelation for *M. catarrhalis* stratified by age. Correction for multiple testing was done using Benjamini-Hochberg correction. **G)** Bar plots showing the number of *S. pneumoniae* (Spn) serotypes that were found co-carried in *lytA* or *piaB*-positive samples. The number of samples versus the number of simultaneously detected serotypes is shown. **H)** Kaplan-Meier survival plot showing the clearance probability of total *S. pneumoniae* (Spn *lytA* in cyan) or of all individual serotypes (red) during the study. Confidence intervals and Chi-square p-value comparing the survival probabilities are indicated. **I)** The odds-ratio (OR) of clearing a *S. pneumoniae* serotype by the next sample, relative to the number of Spn serotypes a child is carrying that day, with single serotype carriage as intercept. The orange dots and bar and whiskers indicate the estimate and standard error of a child clearing a serotype by the next sample, calculated using a binomial mixed model that included child and serotype as random effects and the number of detected serotypes and age as fixed effects. **J)** Density of both *S. pneumoniae* serotypes during dual carriage, relative to total *lytA* density. A serotype within 1 log of *lytA* density is defined as dominant. **K)** Number of times serotypes are observed stratified by the number of carried serotypes in a sample. Black and blue indicate dominant and non-dominant presence of this serotype. Statistical results from binomial mixed effect model including the serotype and number of serotypes as fixed effects and donor as random effect to account for repeated sampling. Only samples with more than 1 detected serotype and only serotypes observed in at least 50 samples were included in the model. **L)** The probability that a serotype is dominant in case of dual carriage, relative to day of acquisition of the new strain. **M)** Example of serotype dynamics in one representative child. The individual datapoints and loess regression are shown. Colour indicate total *S. pneumoniae* density (*lytA* and *piaB*) or serotypes observed in this child. Only serotypes carried at any point by this child are included. **N)** Estimate of clearing a serotype the next day in a binomial mixed effect model including number of carried serotypes, dominance or not and the density of a serotype as fixed effects, with child and serotype as random effects. Point estimate and standard errors are shown. *P<0.05, **P<0.01, ***P<0.001.

To assess the stability of these nasal bacteria over time, we performed autocorrelations for each bacterial species individually, increasing the lag from 1-10 days (**Fig. 3D**). *S. pneumoniae* and *H. influenzae* had very high autocorrelations for up to 3 days, and statistically significant autocorrelation were no longer present after 8 days (**Fig. 3D,E**). For *M. catarrhalis* and particularly *S. aureus*, autocorrelations were less pronounced, although there were statistically significant autocorrelations up to 7 days. Stratifying by age indicated that *M. catarrhalis* stability increased in older children (**Fig. 3F**).

### S. pneumoniae intraspecies dynamics and competition

We next assessed intra-species interactions during nasal carriage. From all *S. pneumoniae lytA*/*piaB*-positive samples, 474 had one detected serotype/group, while 428 samples were positive for multiple serotypes, with up to 6 serotypes/groups detected simultaneously (**Fig. 3G**). For 219 samples, no serotype/group could be identified at all, which is possibly because the panel of serotype-specific assays did not cover all existing serotypes. Although *S. pneumoniae* is traditionally seen as a persistent colonizer, which colonizes for months (*9, 22*), Kaplan-Meier survival curves showed that 51.5% of the carriage episodes were cleared within 17 days at a serotype level (**Fig. 3H**). To understand what was driving clearance of *S. pneumoniae* strains, we used a binomial mixed effects model to predict whether a carried serotype would be cleared by the next sample. Clearance was not associated with age, but strongly associated with the presence of >1 co-carried *S. pneumoniae* serotype/group (**Fig. 3I** and **Fig. S8H**). With 2 serotypes, the probability of clearing a serotype was 3.0x higher compared to not co-carrying another serotype (p=0.0004). This further increased when carrying 3, or 4 or more serotypes/groups (adjusted OR=5.5, p=0.0001). When comparing densities of *S. pneumoniae* serotypes/groups with total *S. pneumoniae lytA* density during co-carriage, the density of one of the carried serotype was typically close to the total *S. pneumoniae* density, while the remaining serotypes/groups had a lower density (**Fig. 3J** and **Fig. S7**). We defined any serotype/group for which the density was within one log_10_ of the *lytA* density as being dominant. The serotypes Spn19A (p=5.6×10^−8^) and Spn10A (p=3.1×10^−5^) were most often dominant (**Fig. 3K**). Spn19A was previously also found to become dominant during mixed serotype inoculation in mice (*23*). Being dominant was not related to being newly acquired or already carried as 60% of the newly acquired serotypes achieved a dominant state by 2 days after acquisition (**Fig. 3L, M**). To understand how dominance was related to the increased risk of clearance during multiple serotype carriage, we refined the clearance model by adding in whether a serotype was dominant or not, as well as the density per serotype (**Fig. 3N**). A dominant serotype was 2.6x less likely to be cleared than a non-dominant serotype (p=0.020), even when correcting for carriage density. Moreover, after correcting for these factors the number of Spn co-carried serotypes was no longer associated with the probability of clearance. Together, this suggests that intra-species competition between *S. pneumoniae* serotypes mediates accelerated clearance of non-dominant strains during co-carriage of multiple serotypes.

### Viral-bacterial interaction dynamics

Viral carriage was common during the study, with on average 1.7 viruses carried per child (range 0-5, **Fig. 4A**). In line with existing literature (*24*), we observed that viral carriage episodes were typically short-lived, with 50% cleared in 9 days (**Fig. 4B**). Bocavirus however could be found in some 1-2 year olds throughout the study period and was thus better at persisting (**Fig. 4A, B**). To understand whether other carried viruses or bacteria were linked to viral clearance, we tested the probability of clearing a virus by the next sample (**Fig. 4C**). Strikingly, *S. aureus* carriage was associated with an increased chance of viral clearance (OR=3.1, p=0.001), while the number of co-carried viruses (OR=0.75, p=0.40) and age (OR=0.98, p=0.86) were not. Thus, *S. aureus* was not only linked to lower probability of acquiring a virus, but also to a greater chance of clearing it. The other way around, viral carriage did not affect *S. aureus* acquisition or clearance (**Fig. S9**). Testing whether *S. aureus* carriage associated with specific viral groups indicated a negative association with carriage of any virus, and of enterovirus and rhinovirus specifically (**Fig. 4D**). There was no significant association with any of the other viral groups, potentially due to limited sample sizes.

**Figure 4.**
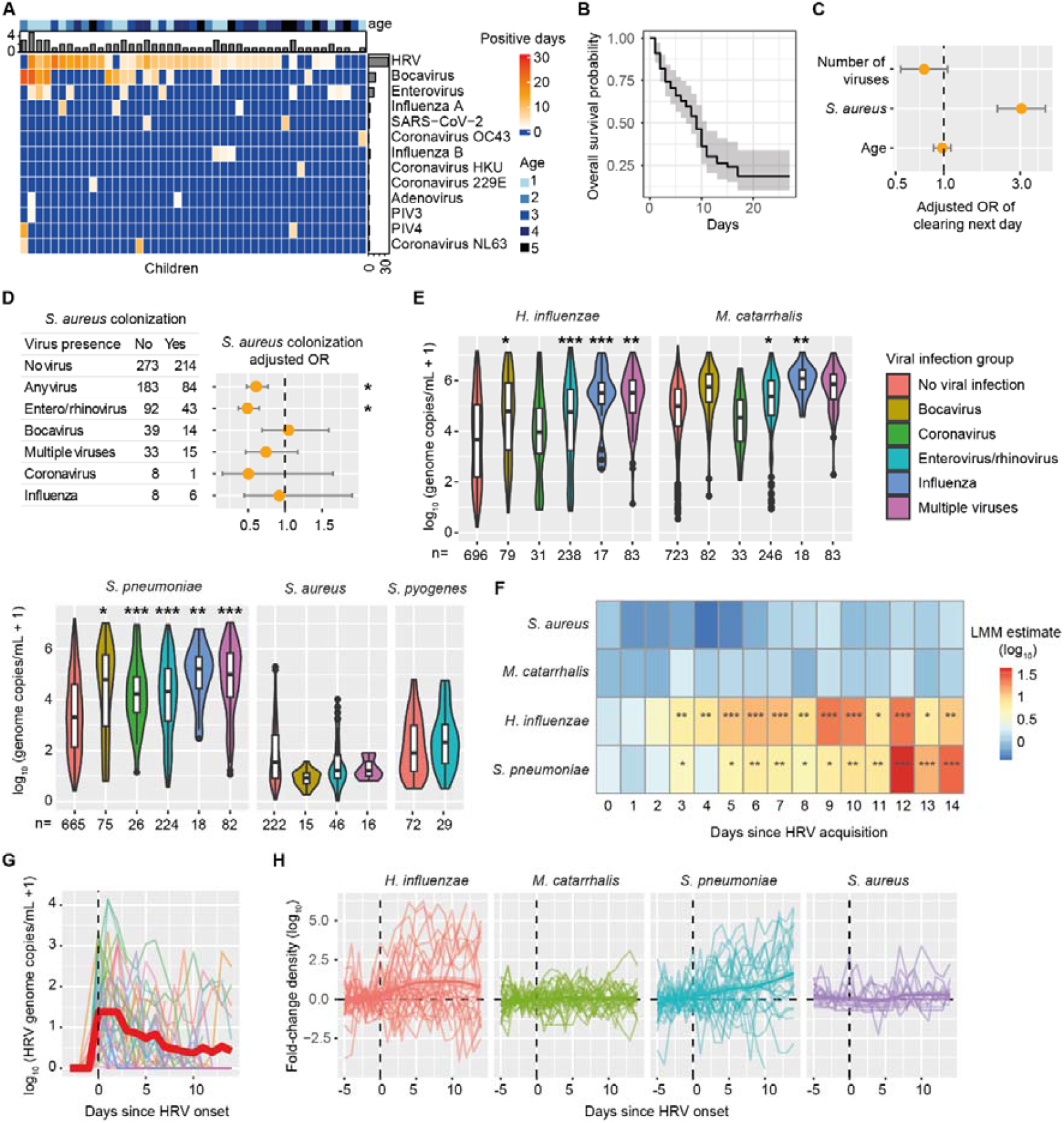
Viral-Bacterial interactions in children. **A)** Heatmap of number of days of viral carriage per child (columns) and virus (rows). Blue indicates a virus was never carried by a child. The colour at top indicates the age per child, ranging from light blue (1 year old) to dark blue (5 years old). The upper bar graph indicates the sum of viruses carried per child. The bar graph on the right indicates the sum of children carrying a virus during the study. **B)** Kaplan-Meier survival plot showing the clearance probability of any virus during the study. Confidence intervals are indicated. **C)** The adjusted odds-ratio (OR) calculated using a binomial mixed model of clearing a virus by the next sample, relative to the number of viruses carried, age of child and presence of *S. aureus* as fixed effects, with child and virus assay as random effects. The orange dots and bar and whiskers indicate the estimate and standard error. **D)** Association between *S. aureus* carriage and viral carriage. Adenovirus and PIV are excluded due to low sample size. Contingency table and adjusted OR based on mixed effect model are shown. Fixed effects are age and viral carriage and child ID as random effect. **E)** Density of bacterial pathogens stratified by the presence of viruses. To increase sample size, viral species were combined into viral groups. Only combinations with at least 10 samples are shown. Correction for multiple testing was done using Benjamini-Hochberg correction. **F)** Heatmap showing results from linear mixed model (LMM) analyzing log-transformed bacterial density relative to days since HRV acquisition, with density 1-3 days prior to viral acquisition combined as baseline. Correction for multiple testing was done using Benjamini-Hochberg correction. **G)** Line plots showing HRV density over time per donor with positive events (colored thin lines), normalised to day of acquisition (dashed line at 0). The thick red line indicates average density per normalised timepoint. **H)** Fold-change plots of bacterial density relative to HRV acquisition day. Individual children and Loess curves are represented. The 3 days prior to HRV acquisition were averaged and used as baseline to calculate the fold-change. *P<0.05, **P<0.01, ***P<0.001.

Viral carriage has been shown to increase density of concurrently carried bacteria. Using a mixed effect model correcting for age and repeated sampling we analysed the effect of concurrent viral carriage on bacterial density, if at least 10 samples with co-carriage of a bacteria and virus pair were present. As expected, *H. influenzae* and *S. pneumoniae* densities were increased in the presence of viral carriage (**Fig. 4E**). In contrast, *S. aureus* and *S. pyogenes* densities were not significantly affected by the presence of a virus. This viral-bacterial effect was dependent on both the virus and bacterium. For example, coronaviruses were not linked to increased density of *H. influenzae*, while influenza was associated with strongly increased bacterial densities.

We next wanted to understand the temporal effects of viral carriage on bacterial density (**Fig. 4F**). For this, we focused on children acquiring HRV, as this was the most frequently carried virus and was strongly associated with increased bacterial density. We therefore aligned samples to the day of HRV acquisition (**Fig. 4G**). For both *H. influenzae* and *S. pneumoniae*, bacterial density increased significantly from 3 days post HRV carriage onset, demonstrating a lag in the effect of viral carriage on bacterial density (**Fig. 4F,H**). For *S. pneumoniae*, HRV was associated with increased density of both pre-existing and newly acquired serotypes (**Fig. S10**), in contrast to findings from studies in adults where pre-existing carriage was less affected (*25, 26*). Densities were still increased at 14 days post rhinovirus acquisition, at which time HRV had already been cleared in 73.7% of the children (**Fig 4G**). In contrast, *S. aureus* and *M. catarrhalis* displayed no significant increase at any day post HRV acquisition, in agreement with the absence of HRV carriage on their densities.

### Symptoms related to microbial dynamics

Through the daily questionnaires, symptom data was provided 1214 times, of which 751 times (61.7%) a respiratory symptom was reported by parents. The most common symptom was snotty nose (n=674, 55.5%), followed by coughing (n=258, 21.2%, **Fig. 5A**). Sneezing (n=39, across 12 children) and ear pain (n=27, across 6 children) were reported less frequently by parents. Fever, defined as a temperature >38°C, was reported in total 11 times (0.9%) in 5 children and was usually self-treated with paracetamol (n=7). Snotty noses were the only symptom that was significantly age related, 82.8% of questionnaires from 1-year-old children reported a snotty nose, which dropped to 31.6% by age 4 (p=4.5×10^−8^, **Fig. 5B**).

**Figure 5.**
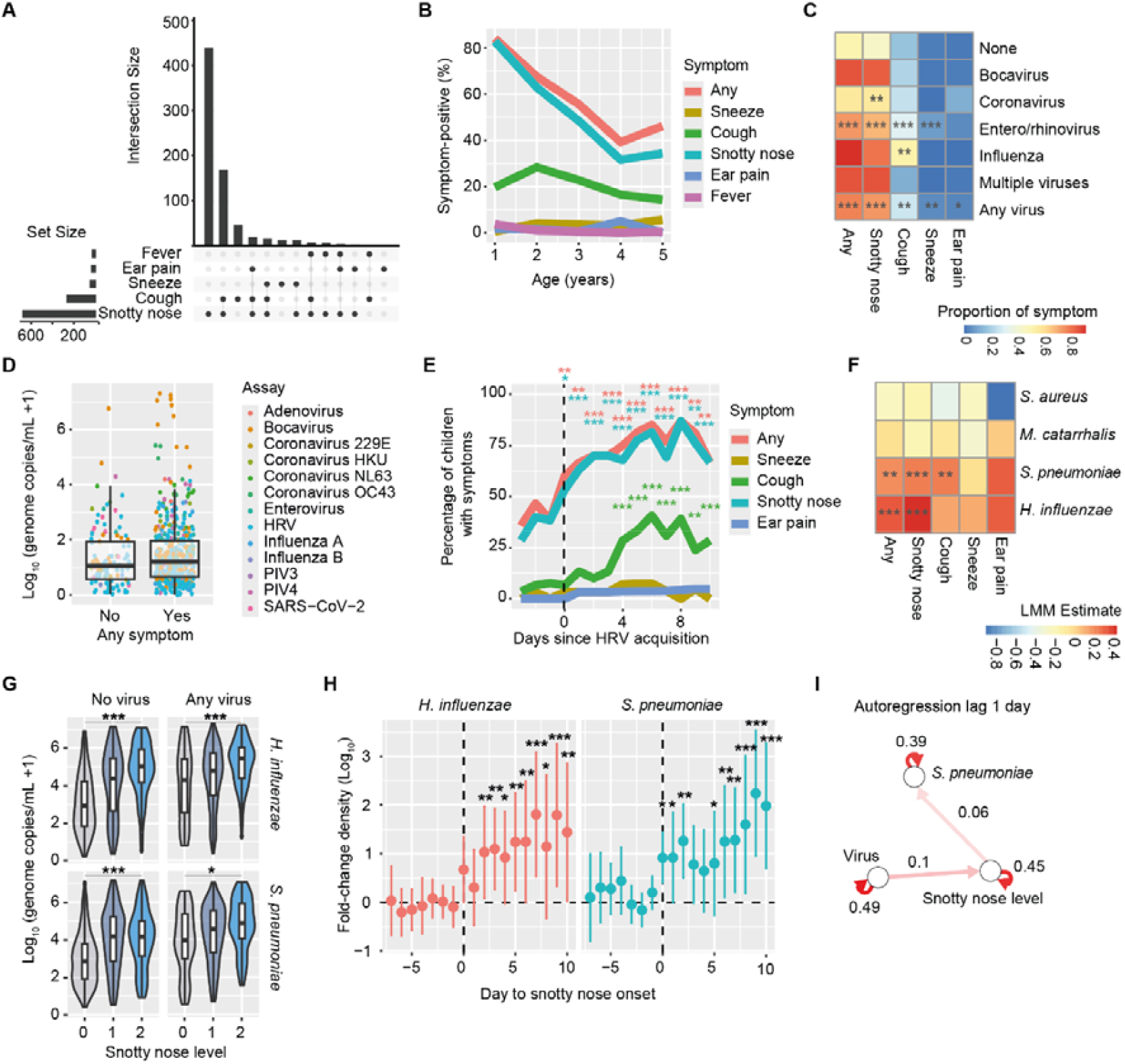
Respiratory symptoms and microbial dynamics in children. **A)** Upset plot showing the total number of times a given symptom was observed for a child, and the intersections of the different symptoms. **B)** Line plots showing the percentage of questionnaires in which a symptom was reported that day, stratified by age. **C)** Heatmap showing the proportion of symptoms, relative to the concurrent presence of viral infections. Statistics are shown from a binomial mixed model that includes age and virus presence as fixed effects and child as random effect. **D)** Boxplots showing viral density stratified by having symptoms or not per sample. Symbols indicate individual samples and symbol colour reflects the virus present. Only samples with virus present are shown. **E)** Line plots showing the percentage of children that shows symptoms, relative to day of rhinovirus (HRV) acquisition. The vertical dashed line indicates the timepoint of viral carriage onset. Statistics are shown from binomial mixed model that includes age and days to carriage onset as fixed effects and child as random effect, with 1-3 days prior to viral acquisition as baseline. **F)** Heatmap showing the estimate of bacterial density on symptom prevalence. Estimates are derived from a single model per symptom, including the bacterial densities and age as fixed effects and child as random effect. ***G)*** *S. pneumoniae* and *H. influenzae* density relative to the amount of snot (0=no snot, 1=a little bit, 2=a lot of snot). Violin and boxplots are shown, stratified by the presence or absence of concurrent viral carriage. **H)** Density of *H. influenzae* and *S. pneumoniae* relative to onset of snotty nose. The fold change compared to baseline (day −3 to −1 relative to symptom onset) is shown. Mean and confidence intervals are shown and dashed lines indicate time of onset and baseline density. Statistics are shown from binomial mixed model that includes age and days to symptom onset as fixed effects and child as random effect, with 1-3 days prior to viral acquisition as baseline. **I)** Temporal network of a multi-level vector autoregression network including viral presence, snotty nose level (0-2) and log-transformed *S. pneumoniae* density. Only significant edges are shown. Arrows indicate directionality of interactions over time. *P<0.05, **P<0.01, ***P<0.001. Multiple testing correction for making separate models per symptom was done with Bonferroni.

Using a mixed effect model combining all viruses and correcting for age and repeated sampling per child demonstrated that viral carriage was significantly associated with all symptoms (**Fig. 5C**). However, viral density was not associated with symptom prevalence, across all viruses and samples (**Fig. 5D**), which is in agreement with earlier findings from SARS-CoV-2 (*27*). Stratifying per virus, rhinovirus/enterovirus was associated with all symptoms except ear pain, and we therefore investigated symptom dynamics relative to HRV carriage onset (**Fig. 5E**). On the day of acquisition, reporting of snotty noses increased significantly, with gradually increasing percentages up to day 8 post HRV acquisition. In contrast, coughing was only found significantly increased from 4 days after first carriage detection onwards. Symptom rates were high in children of all ages after HRV carriage acquisition, although some 4-5 year old children were asymptomatic during HRV carriage, which was not seen in 1-3 year olds (**Fig. S11A**).

Investigating bacterial densities in relation to symptoms showed that *H. influenzae* and *S. pneumoniae* were associated with snotty noses, correcting for age, repeated sampling and other bacteria (**Fig. 5F**). The associations between snotty noses and *S. pneumoniae* (p=6.0×10^−12^) and *H. influenzae* (p<1×10^−16^) carriage densities were independent of viral carriage (**Fig. 5G**). *S. pneumoniae* density was also associated with coughing independently of viral carriage (**Fig. S11B**), while *H. influenzae* was not. Bacterial densities were not significantly linked to sneezing or ear pain. *S. aureus* and *M. catarrhalis* densities were not associated with any symptom (**Fig. 5F**).

We then assessed viral and bacterial carriage dynamics relative to symptom onset. We could identify onset of coughing and snotty noses for 18 and 19 children, respectively. At the day of symptom onset, >50% of children with symptoms carried a virus, while six days prior to snotty nose onset only 7.1% of children carried a virus (**Fig. S11C**). Aligning bacterial carriage density to the day of symptom onset revealed that *S. pneumoniae* density increased about 10-fold on the day snotty noses appeared compared to the 3 days before (p=0.012, **Fig. 5H**). Density continued to increase in the 10 days afterwards to more than 100-fold increases compared to before symptom onset. For *H. influenzae*, kinetics and effect magnitudes were similar, but statistically significant increases started being observed from 2 days post symptom onset. For coughing, viral carriage rates and bacterial densities were less closely related to onset in time than for snotty noses (**Fig. S11C, D**).

To untangle the close link between viral carriage, snotty noses and *S. pneumoniae* or *H. influenzae* density, we employed vector autoregression models with a 1-day lag (**Fig. 5I** and **Fig. S11E**). These models indicated that viral carriage preceded rhinitis, which in turn associated with later increases in bacterial density. In contrast, bacterial densities were not predictive of subsequent increases in rhinitis.

## Discussion

We conducted a unique study with daily nasal sampling to explore microbial dynamics and interactions in young children. This high-frequency approach revealed both expected and novel findings, such as the observation that *M. catarrhalis* density fluctuated with the time of day. While *S. pneumoniae* infection models show significant time-of-day effects on bacterial burden and immune responses, no equivalent studies regarding circadian carriage exist for *M. catarrhalis* in humans or animals to our knowledge (*28*). Daily fluctuations of the gut microbiota have also been observed, which was shown in mouse models to be due to the intestinal epithelial cell clock rather than bacteria-intrinsic, suggesting that host circadian mechanisms in the nasal epithelium might influence *M. catarrhalis* density (*29, 30*).

An unexpected result was the negative association between *S. aureus* and viral carriage. While *S. aureus* is often implicated in secondary pulmonary infections following viral illnesses (*31–33*), our data showed that *S. aureus* carriage associated with a reduced likelihood of acquiring a virus and with accelerated viral clearance. A potential explanation for this is that *S. aureus* induces immune responses with off-target effects (*34*). This suggests a potentially protective role of *S. aureus* carriage against viral infections, in line with the observation that colonization by *S. aureus* very early after birth may be beneficial in protecting against later respiratory infections (*35*), warranting further investigation into the potential underlying mechanisms.

Children experienced on average more than two viral and five *Streptococcus pneumoniae* serotype exposures over the 29-day study period. In the experimental human pneumococcal carriage model, the inoculum of 80,000 colony-forming units is still detectable using nasosorptions in 83% of study participants after 8 hours (*36*). Thus, as some microbes would have been cleared again by time of sampling, it is likely that exposures were even more frequent than what we measured here. Co-exposures were common, unlike what is typically performed in pre-clinical and controlled infection models, highlighting the real-world complexity. Acquisition success following exposure could be linked to age and concurrently carried bacteria and viruses, depending on the encountered microbe. We found that *S. pyogenes* acquisition probability increased with viral presence, aligning with epidemiological trends during and after the COVID-19 pandemic (*37*). However, concurrent viral carriage did not increase *S. pneumoniae* acquisition, unlike what was found in controlled human infection studies in adults (*38*). One potential explanation may be that pre-existing *S. pneumoniae* carriage in children, but not in adults, affected the impact viruses have on bacterial carriage acquisition.

We found that bacterial carriage was highly dynamic as significant auto-correlations in bacterial densities existed for up to one week, with strong autocorrelations only lasting for a few days. Another novel observation was the high turnover of *S. pneumoniae* serotypes, 50% of serotypes were cleared within 2 weeks and replaced by new serotypes. These observations challenge the dogma of bacterial carriage as a stable, persistent process (*9, 22*). Murine studies suggest that existing *S. pneumoniae* strains inhibit the establishment of new carriage via quorum sensing and the production of bactericidal molecules (*9, 39*). Instead, we observed that this intra-species competition was related to accelerated clearance. An additional surprising finding of this study was that the negative association between *S. aureus* and *M. catarrhalis* was only seen when comparing between subjects, but not within individuals. These observations would have been missed in cross-sectional epidemiological studies that report a negative association between these bacteria, as non-included personal confounders may be involved (*40, 41*). For instance, *S. aureus* nasal colonization has been associated with non-secretor Lewis antigen A status, which may also modify susceptibility to other viruses and bacteria (*42*).

The link between viral carriage, bacterial densities and rhinitis has been recognized for a long time, although the directionality of this association has been a subject of debate. On one hand, higher bacterial densities may trigger immune responses that cause symptoms; on the other hand, mucus can be used as a nutrient source of carbohydrates by bacteria including *S. pneumoniae* and *H. influenzae (43)*. Sampling at very high temporal resolution suggests that bacterial density is not driving symptoms but instead is downstream of rhinitis. Rhinovirus carriage acquisition often precipitated this, leading to near-immediate increases in snotty noses and increased bacterial densities a few days later. Increased bacterial densities persisted after the initial viral trigger was cleared again. However, our study cannot exclude that other factors closely associated with mucus production, such as certain host responses, are affected by viral carriage, and that these drive increased bacterial density (*44*).

A limitation of our study is its focus on known pathogenic microbes, potentially missing interactions with other microbial species. Additionally, reliance on parent-collected samples will have introduced variability, although we included a series of quality control measures to address sampling effects, such as temperature logging and inclusion of total 16S detection. Clinician-collected samples at start and end of study were similar to parent-collected samples in neighboring days, further providing confidence of sample quality. Finally, as in any cohort study, findings may not generalize beyond the study population, in this case healthy, young children from a high-income country that go to school or day care. Expanding studies to other populations may provide insights into the increased susceptibility to infection in for example low-income settings (*45*).

In conclusion, our study provides a detailed and novel view of microbial dynamics in young children, revealing complex intra- and inter-species interactions, which confirmed, challenged or clarified existing assumptions. These findings lay the groundwork for future mechanistic, clinical and epidemiological research.

## Data Availability

All data produced in the present study are available upon reasonable request to the authors

https://github.com/spjochems/SAMSAM

## Acknowledgements

Funded by the European Union. Views and opinions expressed are however those of the author(s) only and do not necessarily reflect those of the European Union or the European Research Council Executive Agency. Neither the European Union nor the granting authority can be held responsible for them. This work is supported by an ERC grant (DailySAM, 101075118) and the Gates Foundation (INV-008088) to SPJ. IBC had a fellowship from the São Paulo Research Foundation (FAPESP), Brazil (Process Number #2023/14279-9). Equine arteritis virus for use as internal PCR control was kindly provided by Jessica C. Zevenhoven-Dobbe and Eric J. Snijder of Leiden University Medical Center. We thank Courtney Olwagen for assistance with the microfluidic qPCR method. We thank Sandra Kaamer van Hoegee - Helmus en Jacqueline Zonneveld - van Buyten for assisting in the clinical study and collecting samples. We thank parents and children for their participation in and contribution to the study.

## Competing interests

The authors declare no competing interests

## Author contributions

Study design: DB, KT, MH, SJ Sample collection: EF, MR

Sample processing and analysis: DH, RS, DES, EG, IBC, AK, IV

Data analysis and interpretation: DH, RS, YS, DB, EW, JV, WM, KT, MH, SJ

Writing manuscript: DH, MH, SJ

Supervision: DF, MH, SJ

## Methods

### Ethics Statement

All legal guardians from the children (typically two parents) provided written informed consent. The study was approved by the medical ethical committee METC-LDD under approval number (NL77975-058-21). The study was registered in clinicaltrials.gov (NCT06049537). The here reported outcomes were defined *a priori* in the study protocol. The study was conducted in accordance with the European Statements for Good Clinical Practice. Participants did not receive any financial compensation.

### Study cohort

Between January 2022 and October 2023, we enrolled 46 children into the study. Children were enrolled in all seasons. Potential participants were primarily recruited in the North-Western part of the Netherlands and approached through the municipalities personal records database, inviting parents of young children to contact the research team, but also by putting up posters at out-patient clinics and youth health services. Inclusion criteria were children aged 1-5 years and attending day care or (pre-)school at least 2 (half) days a week, and the ability and willingness of parents to adhere to protocol-specified procedures. Exclusion criteria were history of respiratory tract infections requiring hospitalization, use of antibiotics in the four weeks prior to study enrollment, use of immune-altering medication, and history of severe concomitant disease.

After inclusion, a clinical team member conducted a home visit and collected the first nasosorption sample. A blood and saliva sample were also collected at this visit. The nasosorption procedure was demonstrated to the parents who also received written instructions on how to collect samples. In brief, the synthetic absorptive matrix was inserted fully in the nose, and the same nostril was closed with an index finger for 30 seconds. The first 10 participants were sampled for 60 seconds, which did not significantly affected the study results. Parents were asked to collect samples as much on the same time of day as possible. A daily questionnaire was sent to parents in the evening asking to report on symptoms, medication use, sample collection time and potential sampling issues and activities during the past day. These questionnaires were entered directly by parents into a database (ResearchManager). Parents were asked to store nasosorptions in their home freezer. A USB temperature logger (EL-USB-1, Lascar Electronics) was used to confirm samples remained frozen during the study. At day 14 and 29 (+/− 3 days) additional home visits took place with and collected samples were transported back to the laboratories on dry ice and stored at −70°C until elution. Saliva samples (day 14 and 29; +/− 3 days) and a nasopharyngeal swab (day 29, +/− 3 days) were also collected by a clinical team member during this study visit.

### Nasosorption elutions

Samples were eluted in batches per child, including one mock elution of an untouched nasosorption per child. Nasosorption samples were eluted by adding 100 µL of sterilized elution buffer (PBS + 1% high-grade sterile BSA + 0.05% Triton-X) dropwise on both sides of the absorptive matrix. This run-off was aspirated and re-added to the nasosorption device, until the matrix was visibly saturated. The nasosorptions were then spun at 4,000xg for 10 minutes at 4°C degrees. Total volume per sample was recorded and moved to a sterile Eppendorf tube, and spun at 16,000 x g for 10 minutes at 4°C degrees, to pellet the bacteria. The pellet and half of the supernatant volume was kept at −80°C until nucleic acid extraction. All work was performed in sterile conditions to avoid sample contamination.

### Nucleic acid extraction

All samples were topped up with sterile PBS until 200 µl final volume. Mucous in samples was mechanically homogenized with a pipette tip, and recorded accordingly. MagMAX™ Viral/Pathogen Ultra Nucleic Acid Isolation Kit (Thermo Fisher Scientific, cat. A42356) and the provided instructions were followed and nucleic acid extraction was performed with the Thermo KingFisher Flex 711 Automated Extraction & Purification System. Approximately 70-80 µL elution volume was collected and stored at −80°C until pre-amplification and pathogen detection qPCR.

### Microfluidic qPCR

Targets for 16 viruses, 6 bacterial species, 23 *S. pneumoniae* serotypes/groups and 2 controls (EAV and 16S) were selected for this study (**Table S1** and **Table S2**), based on most prevalent pathogens in children aged 1 to 5 years old in the Netherlands. Microfluidic qPCR protocols were adapted from Olwagen et al (*46*). We tested the microfluidic qPCR by running samples with known positivity for certain bacteria and viruses, assessing *S. pneumoniae* serotypes/groups in *lytA*- and *piaB*-negative samples, comparing sensitivity and specificity relative to conventional *lytA* qPCR, and comparing *lytA* and *piaB* results within samples (**Fig. S4**). Based on this, 19B/F and 18A/B/C were excluded from analyses. For influenza A, all positive samples (Ct<40) were confirmed using a single-plex assay with the same primers/probe, as the pre-amplification of this primer set with other primers led to spurious false-positive results. Samples that could not confirmed with the single-plex as being positive were defined as undetected.

### Pre-amplification

High-throughput nanoliter qPCR reactions require low sample volumes with high nucleic-acid concentration. Therefore, a specific target amplification (STA) was performed. Briefly, in a DNA-free hood, three separated primer pools stocks were prepared containing 180 nM (pool A and B) or 225 nM (pool C) per primer, mixing forward and reverse primers (**Table S2**) in sterile TE buffer (10mM Tris/HCL and 0.1mM EDTA, pH 8). To minimize batch effects, enough volume preamplification primer pools was prepared to run all samples included in this study, then kept at −20°C. On day of preamplification, the required volume of primer pool was collected and TE buffer added to a final concentration of 180 nM (4X for pool A and B) or 225 nM (5X for pool C).

For DNA, per sample, 2.2 µL Preamp Master Mix (Standard Biotools), 2.75 µL 4X primers pool stock A or B (45nM final concentration), and 3.3 µL nuclease-free water were mixed. Then, 7.5 µL was aliquoted in a 96-well PCR plate. Nucleic acid extracted samples were mixed and spun 1000 x g for 30 seconds to pull-down potentially remaining extraction beads. Then 2.5 µL of nucleic acid extracted sample was added to each of the pre-amplification pools of primers. Reactions were amplified in Bio-Rad CFX96 Thermal Cycler with the following condition: 95°C for 2 minutes followed by 14 cycles of denaturation at 95°C for 15 seconds and annealing/extension at 60°C for 4 minutes.

For RNA, 2.75 µL of 5X pooled primer stock (45 nM final concentration), 6.875 µL of 2X RT-PCR buffer and 0.55 µL of 25X AgPath-ID One-Step RT-PCR enzyme mix (ThermoFisher Scientific) and 1.171 µL EAV (1.6-3.2*10^4^ CFU) in nuclease-free water was mixed. Extracted RNA from EAV was used as an internal spike in to assess RT-qPCR efficiency (*47*). Ten µL mix was aliquoted and 2.5 µL of nucleic acid extracted sample was added. The reverse-transcription and pre-amplification were performed with the following cycle program: 45°C for 10 minutes, 95°C for 10 minutes for transcriptase inactivation, then for 14 cycles denaturation at 95°C for 15 seconds and annealing/extension at 60°C for 4 minutes.

Pre-amplified samples were kept in at 4°C overnight and then diluted 1:5, by mixing 5 µL of each preamplification pool (A+B+C) with 10 µL of TE buffer. Pre-amplified samples were then kept at −70 oC until pathogen detection qPCR.

### High-throughput pathogen detection with Biomark HD system

For this study, Standard Biotools 96.96 Dynamic Array™ integrated fluidic circuits (IFC) for Gene Expression were used. Instructions and volumes provided below are per IFC. Per IFC, 90 biological samples and 6 controls were run, including: preamplification negative control, PCR negative control and four dilutions of external synthetic calibrator (gBlock) controls. Per child also an untouched nasosorption was taken along as a mock to demonstrate absence of contamination during laboratory procedures. A total of 17 IFCs were run for all samples in this study.

Target-specific mixes and sample mixes were prepared separately and pipetted accordingly on the IFC on the day of the Biomark run. For the 48 target-specific assay mixes, to minimize batch effects, pre-mixes were prepared for all samples run in this study and kept in a PCR-clean 96-well plate at −20°C. In a DNA-free hood, target-specific assay pre-mix was prepared by mixing forward/reverse primers (18000 nM final concentration, per primer) and probe (5000 nM final concentration) in nuclease-free water to a final volume, preparing 11.5 μL (including 15% overage) per assay target per IFC. Close to the day of the Biomark run, 5 μL target-specific assay pre-mix was mixed with 5 μL 2X Assay Loading Reagent (Standard Biotools) preparing duplicates of each target in a PCR-clean 96-well plate and obtaining a final concentration of 9000 nM forward/reverse primers (per primer) and 2500 nM probe. These target mixes were stored for maximum 3 weeks in a 96-well PCR plate and frozen at −20°C. On the day of Biomark run, assays pre-mixes were thawed, vortexed briefly and pulse-spun before use.

For the sample mixes, on the day of Biomark run, master pre-mix was prepared, sample master pre-mix was prepared in a DNA-free hood by mixing 360 μL of TaqMan Gene Expression (Applied Biosystems) with 36 μL 20X GE Sample Loading Reagent (Standard Biotools) in a 1.5 mL sterile tube and vortexed briefly and pulse-spun. Then, 3.3 μL of sample master pre-mix was aliquoted per well in a sterile PCR plate and 2.7 μL of pre-amplified sample (A+B+C; prepared as described above) added, briefly vortexed and pulse-spun. This is will be referred to as the sample mastermix.

For the Biomark run, manufacturer instructions were followed. Control line fluid was injected into each accumulator on the IFC, and placed in the HX equipment for priming. After priming, 5 μL of each target-specific assay mix (each of the 48 targets in duplicate) and 5 μL each sample master mix was loaded into their respective inlets on the IFC. HX equipment was then used to load the IFC. Afterwards (within 1 hour of loading samples), the IFC was placed in the Biomark HD system for Real-time qPCR using the following thermal cycling conditions: 50°C for 2 min, 70°C for 30 min, 25°C for 10 min, 50°C for 2 min, 96.5°C for 10 min followed by 40 cycles of 96°C for 15 sec, 60°C for 60 sec.

### gBlocks

Synthetic double-stranded DNA (Integrated DNA Technologies), gBlocks, were used as external calibrators as positive control for qPCR, to correct for inter-assay variation, and as a standard curve to calculate gene copies amplified in biological samples (*48*). For each pool, two gBlocks were designed including six to nine targets (**Table S3**). The structure of each gBlock comprised of forward primer sequence, probe sequence and complementary-inverted reverse primer sequence, separated buffer sequences consisting of 5-10 random base pairs (RBP). Each target was separated by buffer sequences consisting of 25 RBP and each gBlock contained buffer sequences of 20-50 RBP at 5’ and 3’. Each individual RBP sequence was unique, and compatibility was tested in silico and validated by qPCR.

### Conventional qPCR

Conventional qPCR was performed for *lytA* using identical primers to validate Biomark results for detection of *S. pneumoniae*. qPCR Master mix was prepared with (per sample), 4.425 µl of nuclease-free water, 0.225 µl of forward and reverse primer (100 µM final concentration, each), 0.125 µl of probe (100 µM final concentration); and 12.5 µl of ⍰TaqMan Universal PCR master mix (Life Technologies). Nucleic acid extracted samples were mixed and spun 1000 x g for 30 seconds to pull-down potentially remaining extraction beads. 2.5 μL of nucleic acid extracted sample and 17.5 µL of master mix were added to each well. Serial dilutions of pneumococcal DNA extract of known concentration was added to the plates and used as a standard curve to deduct number of copies in each sample. The plates were run with a thermocycler (Applied Bioscience QuantStudio 7 Flex) with the following amplification conditions: 50 oC for 2 minutes, 95°C for 10 min; followed by 40 cycles of 95°C for 15 s and 60°C for 1 min.

### Analysis

Concentrations from bacterial and viral targets were determined using the Fluidigm Real-Time PCR Analysis software (v4.8.1) using a standard curve based on gBlocks. All assays were run in duplicate. An average concentration was determined across duplicates and concentration of undetected assays were set to 0. The threshold for positivity was set at Ct=40. When one of two duplicates for an assay/sample combination were positive, we defined this as that microbe being detected in the sample. Using this threshold, we defined carriage events as an assay being detected at least twice in a 5-day period. This allows for up to three negative samples in between positive samples, as low density carriage around the limit of detection could sometimes be undetectable for several days in a row (**Fig. S6** and **Fig. S7**). For this reason, we defined exposure events as the first occurrence of a pathogen after not having been detected for at least four days. And we defined clearance as having a negative PCR after being carrier, with no more positive PCRs observed in the coming three days. In line with this, we defined symptom onset as not having had a symptom for at least three days and then having it for at least two days in a row. Data analysis was performed in R (v4.4.2), using RStudio (v2024.09). As repeated samples were taken in the study, mixed effects models were used for analysis where the study ID of the participant was added as a random effect using the “lme4” package (v1.1-35.5) and “lmerTest” package (v3.1-3). Vector autoregression modelling was performed using the “mlVAR” package (v0.5.2). Correction for multiple testing was performed using Benjamini-Hochberg and results with p<0.05 were considered significant.

## Code availability

Data analysis code is deposited on https://github.com/spjochems/SAMSAM

## Data availability

Data can be requested from the corresponding author. An MTA will be required to ensure compliance with ethically approved protocol and privacy regulations prior to transfer of data.

## Notes

### Competing Interest Statement

The authors have declared no competing interest.

### Author Declarations

The Medical Ethics Committee Leiden The Hague Delft of the Leiden University Medical Centre gave ethical approval for this work

### Summary of Updates

Influenza A results from Biomark were confirmed with single-assay conventional PCR.

