## Supplementary Figures and Tables for "High-resolution characterization of nasal microbial dynamics in young children"

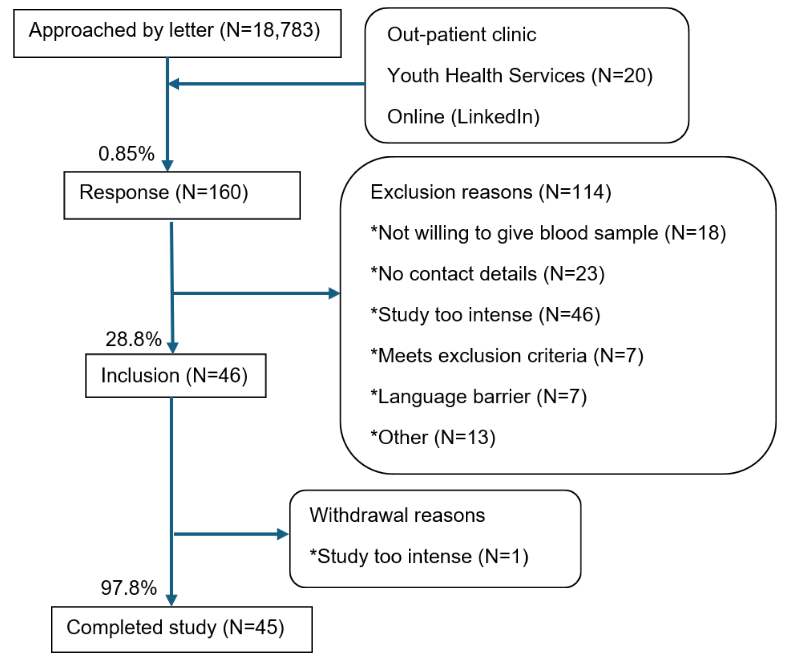

**Figure S1. Study Flow chart.** Numbers of children who were contacted, informed of the study, included in the study and that completed the study are shown. Reasons for not being included are depicted on the right.

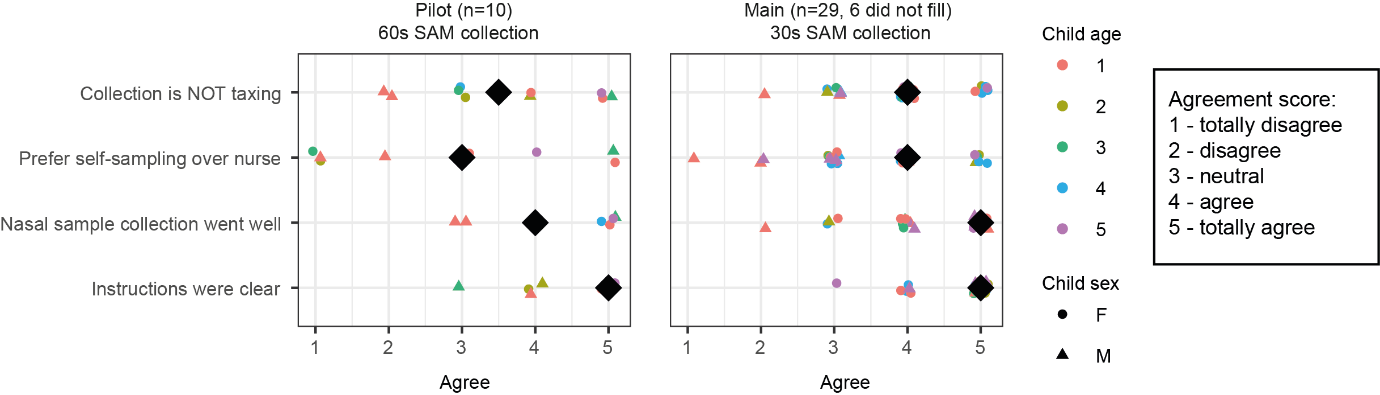

**Figure S2. Study evaluation questionnaire.** Evaluation questionnaires were filled by parents at the end of study. Individual responses and median results (black diamonds) are indicated. Colour and shape indicate age and sex of child, respectively. Evaluation forms are stratified by pilot (left, 10 children) and main (right, 29 children, evaluation questionnaire was not filled in for 6 participants). After the pilot, the nasosorption sampling duration was reduced from 60 seconds to 30 seconds. Parents from the one child that withdrew from the study did not fill out a questionnaire.

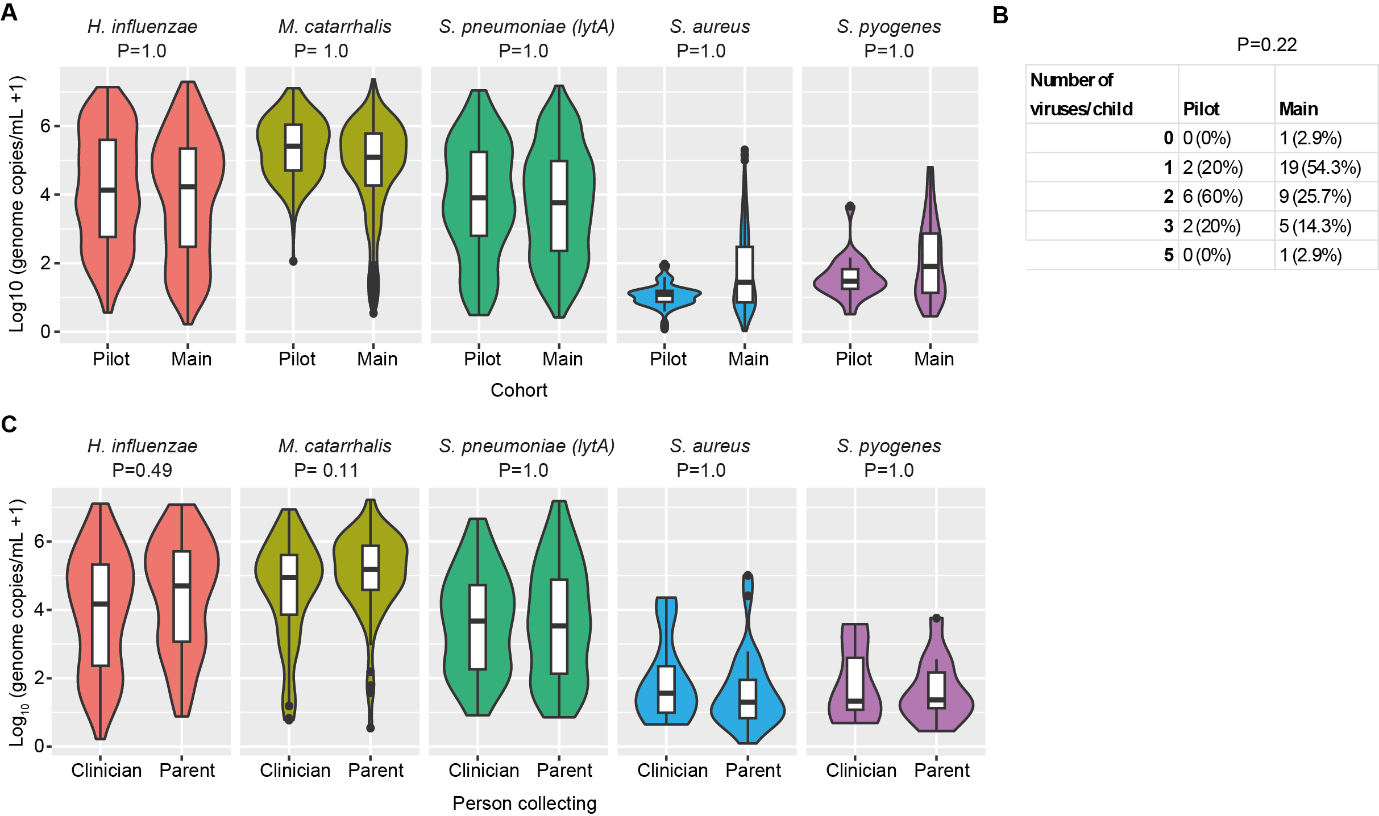

**Figure S3. Nasosorption sampling quality control. A)**. Density of bacteria in samples collected during the pilot cohort (n=10 children, sampling duration 60 seconds) or main cohort (n=35 children, sampling duration 30 seconds). Statistical results from linear mixed model including cohort and age as fixed effect and child ID as random effect. **B)** Table showing the number (and percentages) of viruses detected per child in pilot and main cohort. P-value from Fisher exact test is depicted above the contingency table. **C)** Density of bacteria in samples collected by clinicians, and adjacent samples self-collected by parents. Statistical results from linear mixed model including person collecting as fixed effect and child ID as random effect.

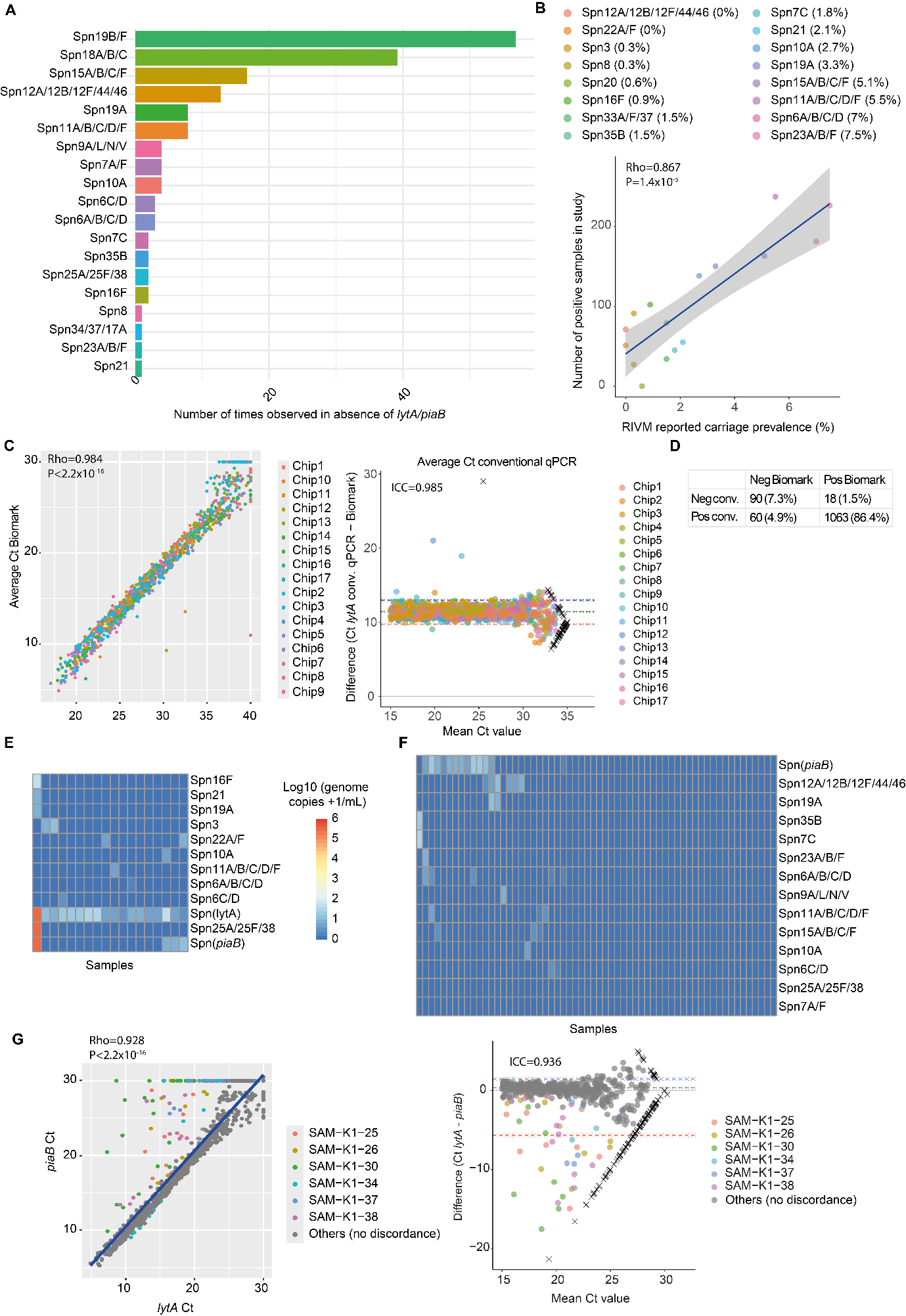

**Figure S4. Biomark sensitivity and specificity.** **A)** Numbers of samples for which a *S. pneumoniae* (Spn) serotype/group was detected, while *lytA* and *piaB* were both negative, indicating a potential lack of specificity. **B)** Correlation plot showing association (Pearson rho=0.867; p=1.4*10^-5^) between the total number of positive samples per serotype detected within this study and the population-level serotype prevalence during the same period, as reported by the Dutch National Institute for Public Health and the Environment (RIVM). Each symbol is one serotype/group, with legend showing the serotype/group and the RIVM reported carriage prevalence. **C)** Correlation plot (left) and Bland-Altman plot (right) of *lytA* Ct values measured by conventional qPCR and Biomark, using the average of two technical duplicates for each method. Pearson rho, p-value and intra-class correlation coefficient (ICC) are depicted. Negative samples were set to Ct=40 for conventional qPCR and Ct=30 for Biomark. **D)** Table showing *lytA* positivity of samples for conventional qPCR and Biomark. Both absolute numbers and percentages are indicated. **E)** Heatmaps of samples positive for *lytA* by Biomark and negative by conventional qPCR, showing all Spn assays that were positive for at least one of these samples. **F)** Heatmaps of samples negative for *lytA* by Biomark and positive by conventional qPCR, showing only Spn assays that were positive for at least one of these samples. **G)** Correlation plot (left) and Bland-Altman plot (right) comparing *piaB* and *lytA* values. Ct values are shown for all samples using the average of two technical duplicates per assay. Colours of symbols indicate children from which the two assays showed discordant results between *piaB* and *lytA*. All other children are depicted in grey. Pearson rho, p-value and intra-class correlation coefficient (ICC) are depicted.

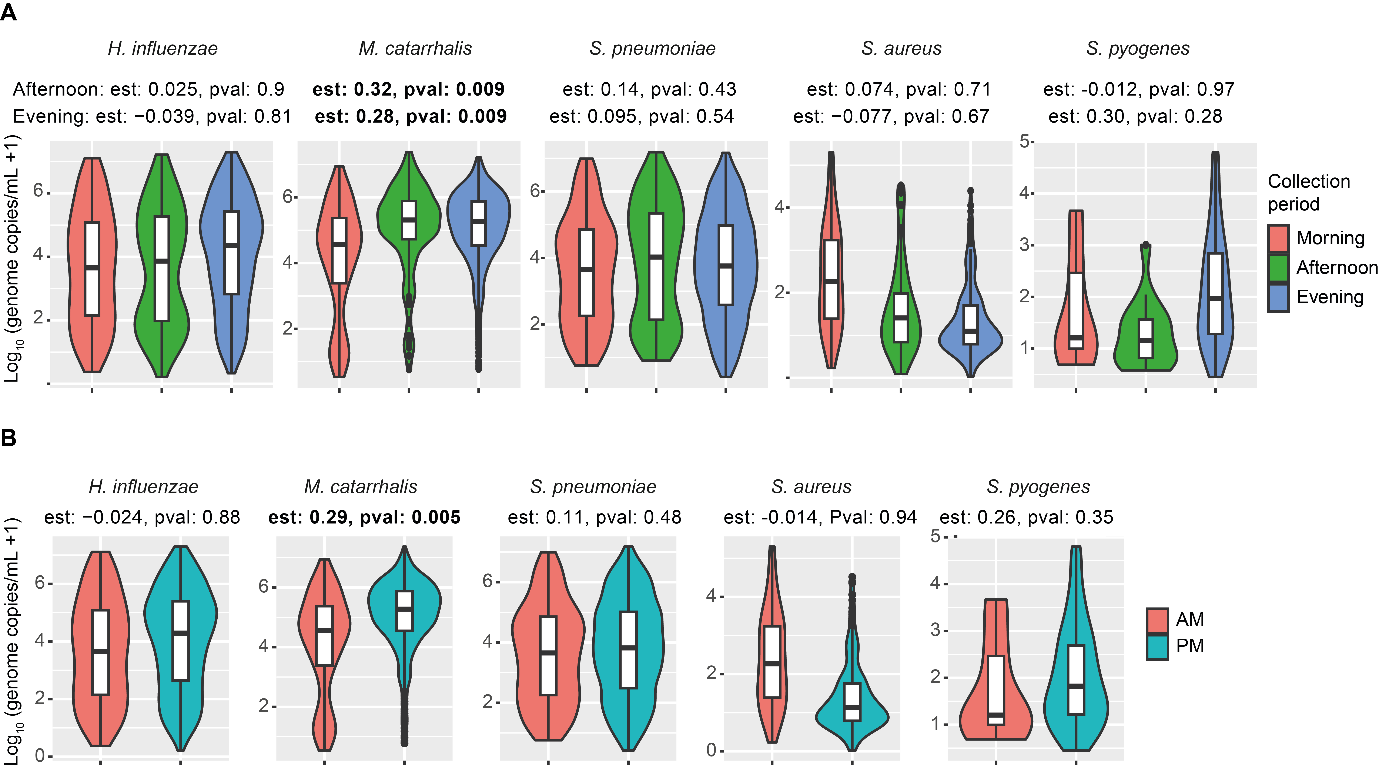

**Figure S5. Time of day for sample collection effect on bacterial density. A)** Density of 5 bacteria in morning (before 12pm), afternoon (12-6pm), or evening (after 6pm) are shown. All collected samples with reported time of collection are shown, excluding bacteria that are undetectable. Violin and boxplots are shown. Statistical results from linear mixed model including child ID as random effect and collection period as fixed effect estimate (Est) are shown, using morning (AM) as base level. **B)** Same data as for A), but combining afternoon and evening samples into PM.

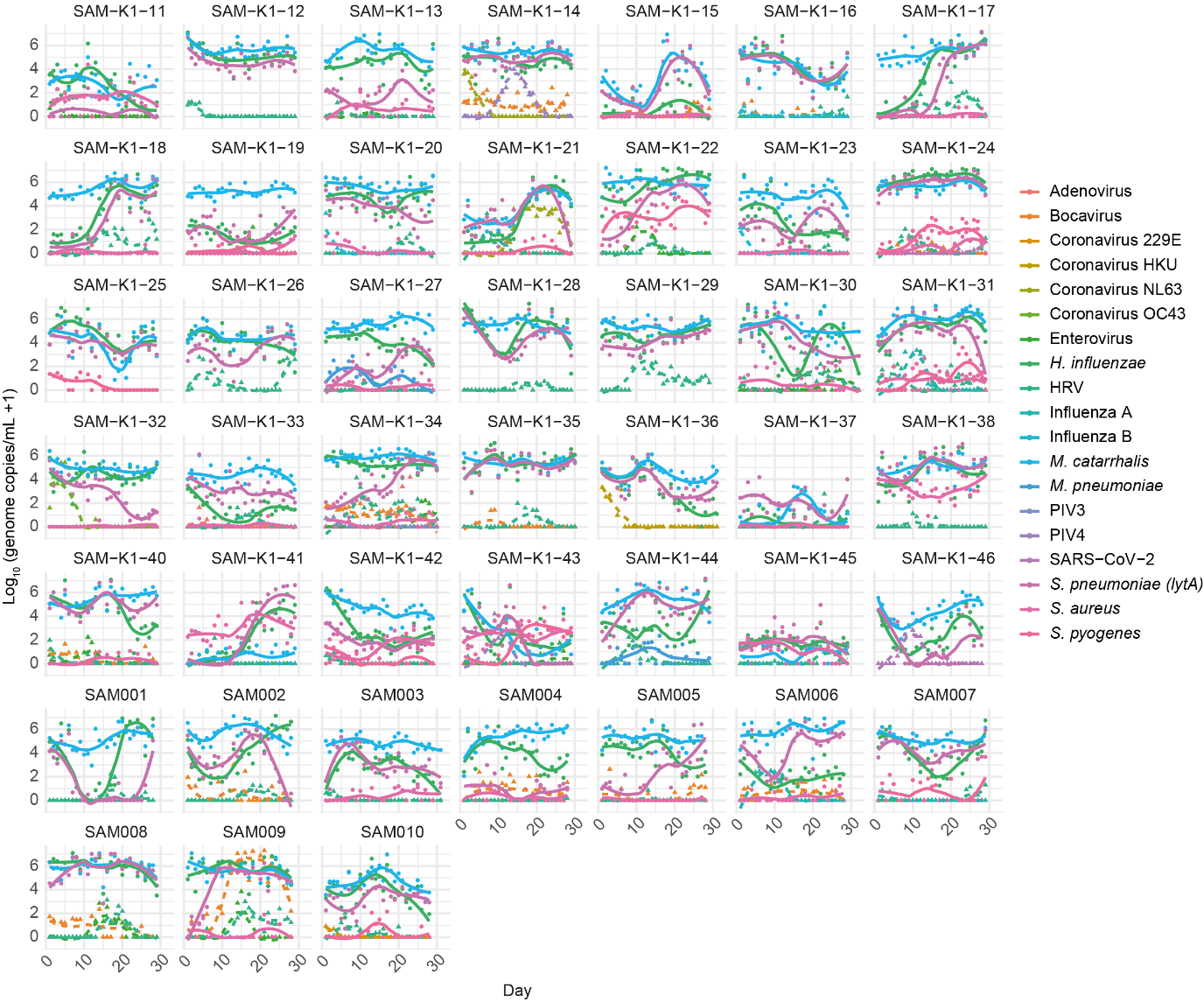

**Figure S6. Bacterial and viral carriage dynamics.** Longitudinal analysis of bacterial (continuous lines and circles) and viral (dashed lines and triangles) carriage densities for all children in the study. Individual points and loess curves are shown. Per child only assays that were carried at any point are shown, with carriage defined as having at least two positive samples in a five-day period.

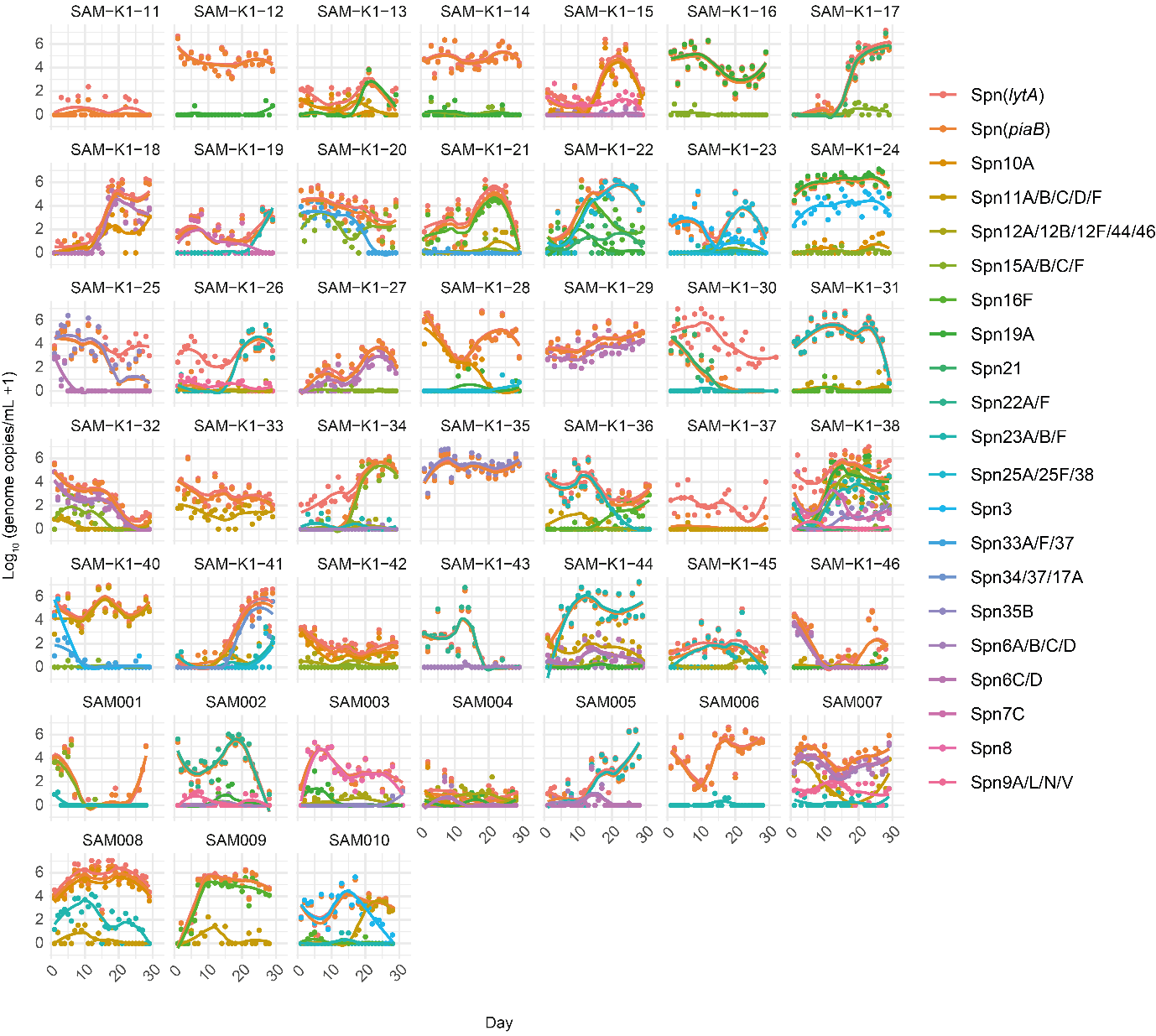

**Figure S7. *S. pneumoniae* serotype carriage dynamics.** Longitudinal analysis of *S. pneumoniae* (Spn) serotype carriage densities for all children in the study. Individual points and loess curves are shown. Per child only assays that were carried at any point are shown, with carriage defined as having at least two positive samples in a five-day period. *lytA* and *piaB* detect *S. pneumoniae* independently of serotype.

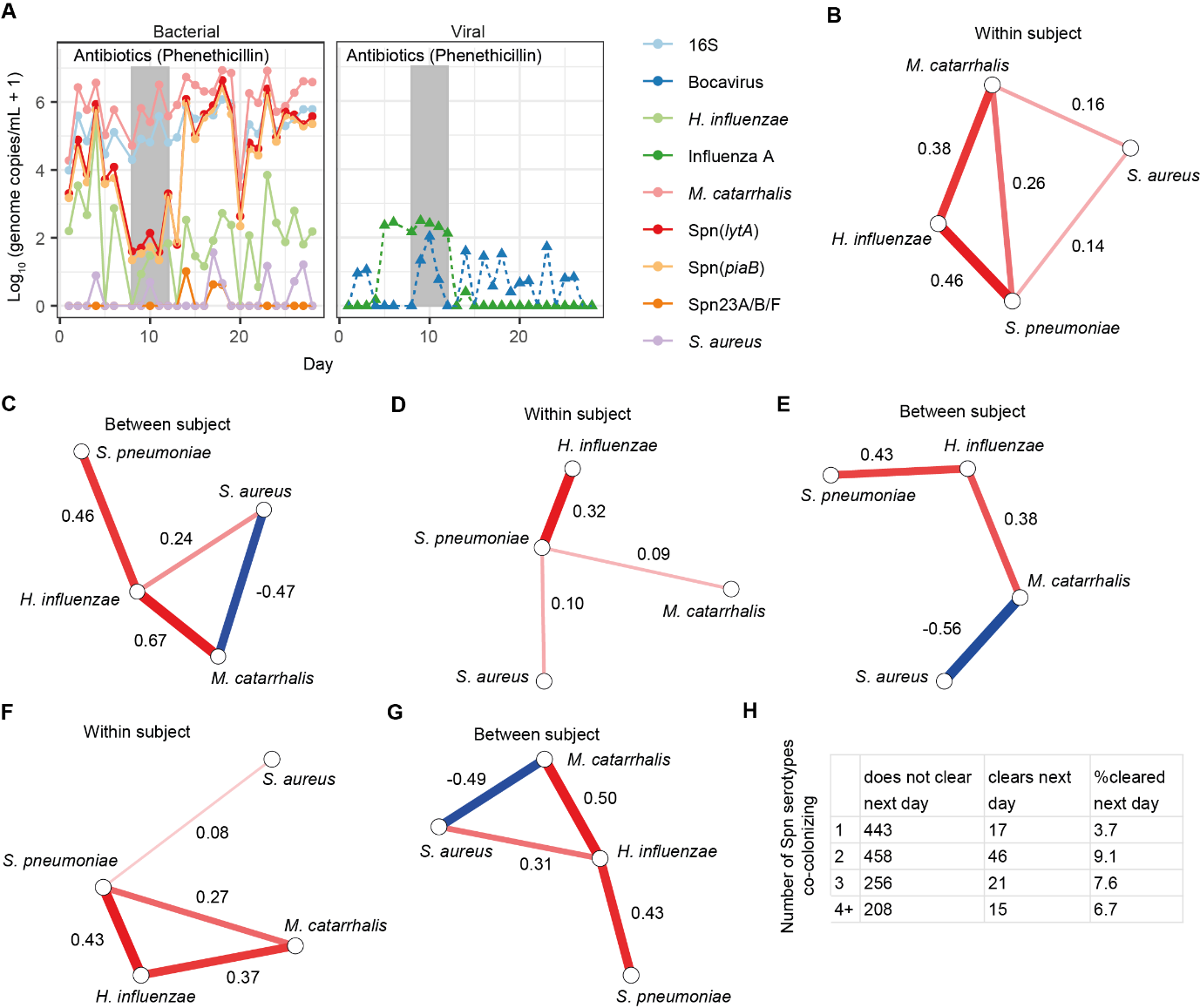

**Figure S8. Bacterial interactions and carriage dynamics in children. A)** Line plots of one child who received antibiotics treatment (grey rectangle) during the study. All bacterial (left) or viral (right) assays that were carried by this child are depicted. **B)** Multi-level vector autoregression modelling showing within subject and **C)** between subject partial correlations between bacterial species. Loess estimates of bacterial densities were used to correct for potential sampling effects. Red and blue lines indicate positive and negative associations, respectively, with only lines with P<0.05 depicted. **D)** Multi-level vector autoregression modelling showing within subject and **E)** between subject partial correlations between bacterial species. Bacterial densities were normalized for total 16S content to correct for potential sampling effects. Red and blue lines indicate positive and negative associations, respectively, with only lines with P<0.05 depicted. **F)** Multi-level vector autoregression modelling showing within subject and **G)** between subject partial correlations between bacterial species, for samples where there was no concurrent viral presence detected. Red and blue lines indicate positive and negative associations, respectively, with only lines with P<0.05 depicted. **H)** Table showing the number of times a *S. pneumoniae* (Spn) carriage is cleared or not by the next sample, stratified by the number of co-carried Spn serotypes.

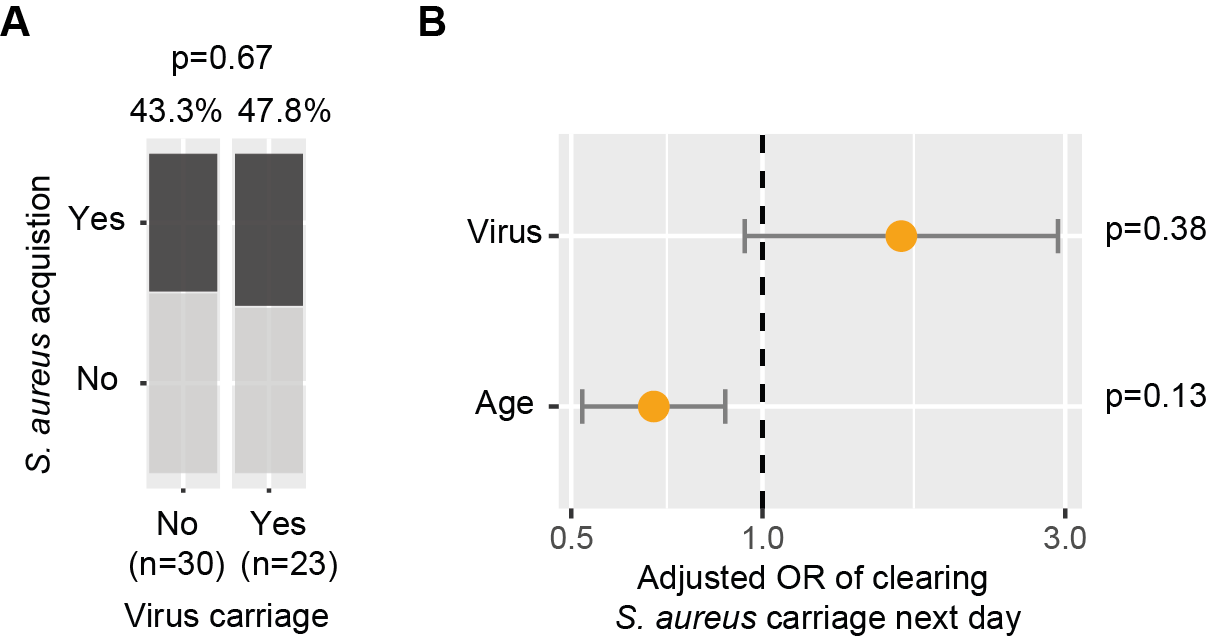

**Figure S9. Viral carriage and *S. aureus* acquisition or clearance.** **A)** Acquisition success of *S. aureus* of children with concurrent viral infection or not. Percentages of acquisition and p-value of binomial mixed effects model, including viral carriage as fixed effect and child ID as random effects, are depicted. **B)** The adjusted odds-ratio (OR) calculated using a binomial mixed model of clearing *S. aureus* carriage by the next sample, relative to viruses carried or age of child as fixed effects, with child as random effect. The orange dots and bar and whiskers indicate the estimate and standard error. P-values of the predictor variables are depicted.

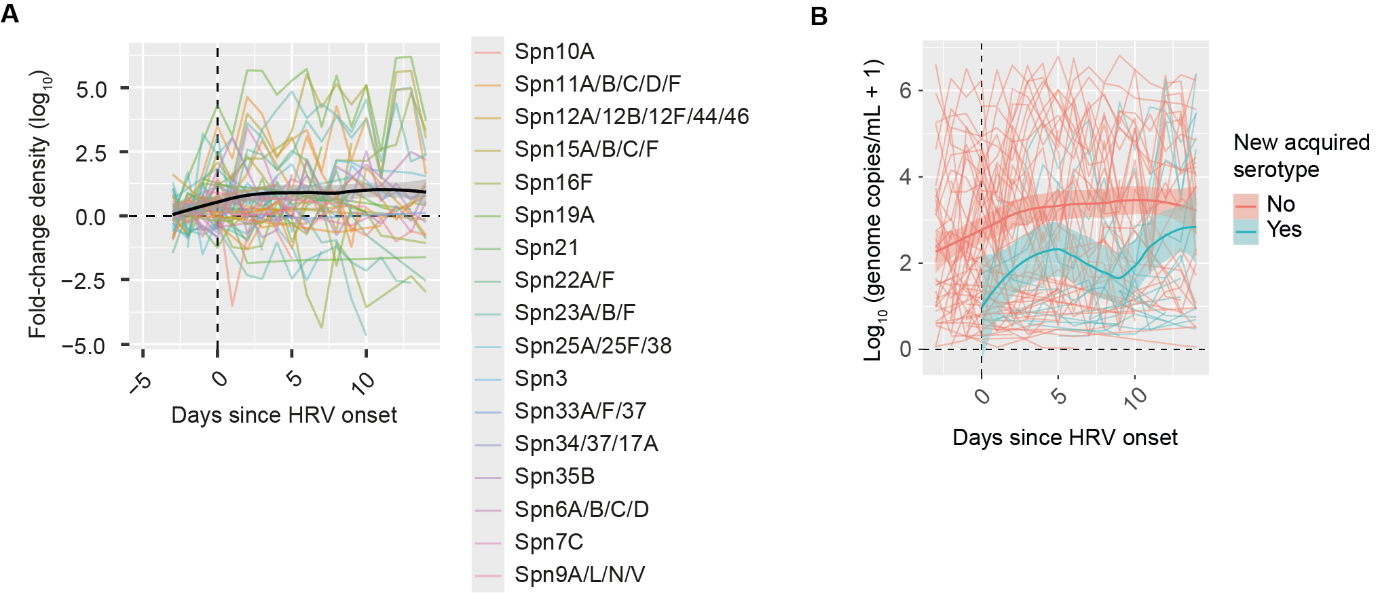

**Figure S10. Rhinovirus carriage and *S. pneumoniae (Spn)* serotype bacterial density.** **A)** Line plots representing fold-change (FC) of bacterial density per *S. pneumoniae* serotype over time, aligned to the day of human rhinovirus (HRV) onset (vertical dashed line at 0). The bacterial density of 3 days prior to HRV acquisition were averaged and used as baseline to calculate the fold-change. Line colours indicate Spn serotypes/groups. The black line depicts the Loess curve across all samples. Only serotypes carried before viral onset are included. **B)** Line plots showing absolute *Spn* density over time. Data is coloured by whether *S. pneumoniae* was detected already pre-HRV acquisition (red) or not (blue). Individual lines and Loess curves are represented.

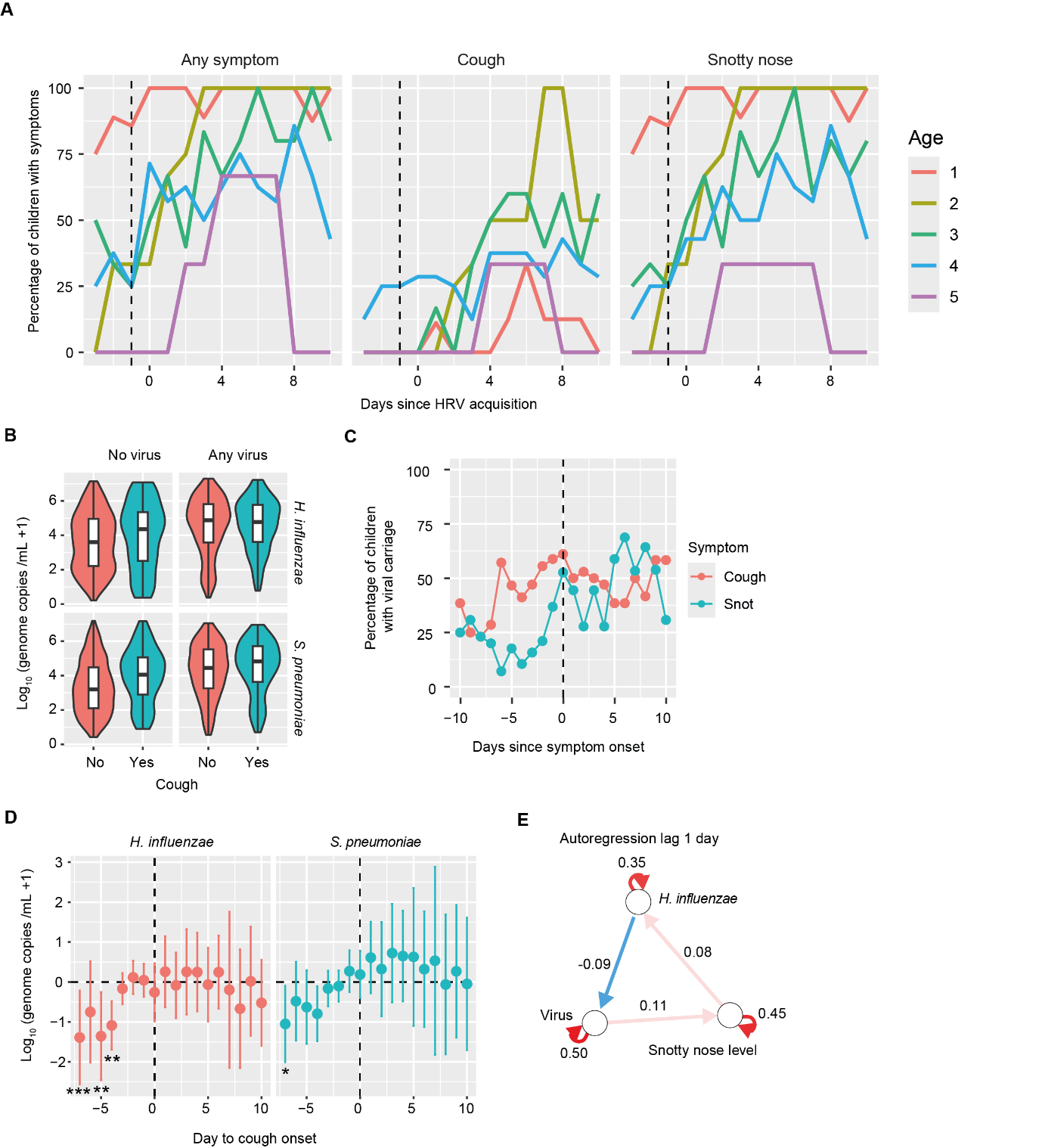

**Figure S11. Respiratory symptoms over time.** **A)** Line plots showing the percentage of children that shows symptoms, relative to the day of rhinovirus (HRV) acquisition. The facets indicate the type of symptom and the colour the age group. The vertical dashed line indicates the last timepoint before infection is observed. N = 9, 4, 6, 8, 3 for the ages 1-5, respectively. **B)** *S. pneumoniae* and *H. influenzae* density relative to coughing. Violin and boxplots are shown, stratified by the presence or absence of concurrent viral carriage. **C)** The percentage of children with viral carriage relative to the first appearance of a snotty nose or coughing. Onset of symptom is defined by not having had a symptom in the three days prior and then having it for at least two days. **D)** Density of *H. influenzae* and *S. pneumoniae* relative to onset of coughing. The fold change compared to baseline (day -3 to -1 relative to symptom onset) is shown. Mean and confidence intervals are shown and dashed lines indicate time of onset and baseline density. Statistics are shown from binomial mixed model that includes age and days to symptom onset as fixed effects and child as random effect, with 1-3 days prior to viral acquisition as baseline. *P<0.05, **P<0.01, ***P<0.001. **E)** Temporal network of a multi-level vector autoregression network including viral presence, snotty nose level (0-2) and *H. influenzae* density. Only significant edges are shown. Arrows indicate directionality of interactions over time.

**Table S1.** Targets used for qPCR detection of bacterial, virus, *Streptococcus pneumoniae* (Spn) serotyping (ST) or serogrouping (SG) and controls.

**Bacteria**

*Haemophilus influenzae*

*Moraxella catarrhalis*

*Mycoplasma pneumoniae*

*Staphylococcus aureus*

*Streptococcus pneumoniae*

*Streptococcus pyogenes*

**Spn ST/SG**

3

6A/B/C/D

6C/D

7A/F

7C

8

9A/L/N/V

10A

11A/B/C/D/F

12A/12B/12F/44/46

15A/B/C/F

16F

19A

20

21

22A/F

23A/B/F

25A/25F/38

33A/F/37

34/37/17A

35B

**Virus**

SARS-CoV-2

Influenza virus A

Influenza virus B

Respiratory syncytial virus A

Respiratory syncytial virus B

Human rhinovirus

Coronavirus 229E

Coronavirus NL63

Coronavirus OC43

Coronavirus HKU

Parainfluenza virus 1

Parainfluenza virus 2

Parainfluenza virus 3

Parainfluenza virus 4

Enterovirus

Adenovirus

Bocavirus

**Controls**

16S

Equine arteritis virus

**Table S2.** Sequences from primers and probes used for pathogen qPCR detection, Streptococcus pneumoniae (Spn) serotyping (ST) or serogrouping (SG). For preamplification of samples, primers (excluding 16S) were combined in three pools. [+] indicates locked nucleotides to increase melting temperature. Spn18A/B/C and 19B/F were excluded from analysis due to lack of specificity.

| **Assay** | **Pool** | **Primer/Probe** | **Sequence (5’- 3’)** | **Source** |
| --- | --- | --- | --- | --- |
| 16S | - | 16S-Fwd | TCCTACGGGAGGCAGCAGT | Nadkarni *et al*, 2002 (*1*) |
|  |  | 16S-Rev | GGACTACCAGGGTATCTAATCCTGTT |  |
|  |  | 16S-Probe | (FAM)-CGTATTACCGCGGCTGCTGGCAC-(BHQ) |  |
| Adenovirus | A | ADV-Fwd1 | TTTGAGGTGGAYCCMATGGA | van de Pol et al, 2007 (*2*) |
|  |  | ADV-Fwd2 | TTTGAGGTYGAYCCCATGGA |  |
|  |  | ADV-Rev1 | AGAASGGSGTRCGCAGGTA |  |
|  |  | ADV-Rev2 | AGAASGGTGTRCGCAGATA |  |
|  |  | ADV-Probe1 | (FAM)-ACCACGTCGAAAACTTCGAA-(MGB) |  |
|  |  | ADV-Probe2 | (FAM)-ACCACGTCGAAAACTTCAAA-(MGB) |  |
|  |  | ADV-Probe3 | (FAM)-ACACCGCGGCGTCA-(MGB) |  |
| Bocavirus | B | BocaV-Fwd | GGAAGAGACACTGGCAGACAA | Allander *et al*, 2007 (*3*) |
|  |  | BocaV-Rev | GGGTGTTCCTGATGATATGAGC |  |
|  |  | BocaV-Probe | (FAM)-CTGCGGCTCCTGCTCCTGTGAT-(BHQ) |  |
| Coronavirus 229E | C | HCoV-229E-Fwd | ACTTTGTCAGTTCGTATGCTAAAC | Zhao *et al*, 2022 (*4*) |
|  |  | HCoV-229E-Rev | TTGTCAGAACATTGGCATTAACA |  |
|  |  | HCoV-229E-Probe | (FAM)-ATTATCAGCTTATGACTTGGCGTGT-(BHQ) |  |
| Coronavirus HKU | C | CoV_HKU-Fwd | AGTTCCCATTGCTTTCGGAGTA | Cui *et al*, 2011 (*5*) |
|  |  | CoV_HKU-Rev | CCGGCTGTGTCTATACCAATATCC |  |
|  |  | CoV_HKU-Probe | (FAM)-CCCCTTCTGAAGCAA-(MGB) |  |
| Coronavirus NL63 | C | CoV_NL63-Fwd | GCGTGTTCCTACCAGAGAGGA | Loens *et al*, 2012 (*6*) |
|  |  | CoV_NL63-Rev | GCTGTGGAAAACCTTTGGCA |  |
|  |  | CoV_NL63-Probe | (FAM)-ATGTTATTCAGTGCTTTGGTCCTCGTGAT-(BHQ) |  |
| Coronavirus OC43 | C | CoV_OC43-Fwd | CGATGAGGCTATTCCGACTAGGT | Loens *et al*, 2012 (*6*) |
|  |  | CoV_OC43-Rev | CCTTCCTGAGCCTTCAATATAGTAACC |  |
|  |  | CoV_OC43-Probe | (FAM)-TCCGCCTGGCACGGTACTCCCT-(BHQ) |  |
| Enterovirus | C | EnteroV-Fwd | CCCTGAATGCGGCTAATCC | Wolffs et al, 2011 (*7*) |
|  |  | EnteroV-Rev | ATTGTCACCATAAGCAGCCA |  |
|  |  | EnteroV-Probe | (FAM)-AACCGACTACTTTGGGTGTCCGTGTTTC-(BHQ) |  |
| Equine arteritis virus | C | EAV-Fwd | GGCGACAGCCTACAAGCTACA | Balasuriya *et al*, 2002 (*8*) |
|  |  | EAV-Rev | CGGCATCTGCAGTGAGTGA |  |
|  |  | EAV-Probe | (FAM)-TTGCGGACCCGCATCTGACCAA-(BHQ) |  |
| *Haemophilus influenzae* | A | IgA1-Fwd | CAAAATTGCCAAGATTAAATGCTT | Olwagen *et al*, 2018 (*9*) |
|  |  | IgA1-Rev | TGCTCGCCATACTGCACAA |  |
|  |  | IgA1-Probe | (FAM)-C[+C][+T]G[+C]GGT[+T]AAAC[+C]-(BHQ) |  |
| Human rhinovirus | C | HRV-Fwd | CY[+A]GCC[+T]GCGTGGC | Lu *et al*, 2008 (*10*) |
|  |  | HRV-Rev | GAAACACGGACACCCAAAGTA |  |
|  |  | HRV-Probe | (FAM)-TCCTCCGGCCCCTGAATGYGGC-(BHQ) |  |
| Influenza virus A | C | FluA-Fwd | AAGACCAATCCTGTCACCTCTGA | Loens *et al*, 2012 (*6*) |
|  |  | FluA-Rev | CAAAGCGTCTACGCTGCAGTCC |  |
|  |  | FluA-Probe | (FAM)-TTTGTGTTCACGCTCACCGTGCC-(BHQ) |  |
| Influenza virus B | C | FluB-Fwd | AAATACGGTGGATTAAACAAAAGCAA | Loens *et al*, 2012 (*6*) |
|  |  | FluB-Rev | CCAGCAATAGCTCCGAAGAAA |  |
|  |  | FluB-Probe | (FAM)-CACCCATATTGGGCAATTTCCTATGGC-(BHQ) |  |
| *Moraxella catarrhalis* | A | MCAT-Fwd | CGTGTTGACCGTTTTGACTTT | Dunne *et al*, 2012 (*11*) |
|  |  | MCAT-Rev | TAGATTAGGTTACCGCTGA[+C]G |  |
|  |  | MCAT-Probe | (FAM)-ACCGACATCAACCCAAGCTTTGG-(BHQ) |  |
| *Mycoplasma pneumoniae* | A | MPn-Fwd | GGTCAATCTGGCGTGGATCT | Loens *et al*, 2012 (*6*) |
|  |  | MPn-Rev | TGGTAACTGCCCCACAAGC |  |
|  |  | MPn-Probe | (FAM)-TCCCCCGTTGAAAAAGTGAGTGGGT-(BHQ) |  |
| Parainfluenza virus 1 | C | PIV1-Fwd | TGATTTAAACCCGGTAATTTCTCAT | van de Pol *et al*, 2007 (*2*) |
|  |  | PIV1-Rev | CCTTGTTCCTGCAGCTATTACAGA |  |
|  |  | PIV1-Probe | (FAM)-ACGACAACAGGAAATC-(MGB) |  |
| Parainfluenza virus 2 | C | PIV2-Fwd | AGGACTATGAAAACCATTTACCTAAGTGA | van de Pol *et al*, 2007 (*2*) |
|  |  | PIV2-Rev | AAGCAAGTCTCAGTTCAGCTAGATCA |  |
|  |  | PIV2-Probe | (FAM)-ATCAATCGCAAAAGCTGTTCAGTCACTGCTATAC-(BHQ) |  |
| Parainfluenza virus 3 | C | PIV3-Fwd | TGATGAAAGATCAGATTA[+T]GCAT | Loens *et al*, 2012 (*6*) |
|  |  | PIV3-Rev | CCGGGACACCCAGTTGTG |  |
|  |  | PIV3-Probe | (FAM)-TGGACCAGGGATATACTACAAAGGCAAAATAATATTTCTC-(BHQ) |  |
| Parainfluenza virus 4 | C | PIV4-Fwd | CAAAYGATCCACAGCAAAGATTC | van de Pol *et al*, 2007 (*2*) |
|  |  | PIV4-Rev | ATGTGGCCTGTAAGGAAAGCA |  |
|  |  | PIV4-Probe | (FAM)-GTATCATCATCTGCCAAATCGGCAATTAAACA-(BHQ) |  |
| SARS-CoV-2 | C | SCOV2-Fwd | CTGCAGATTTGGATGATTTCTCC | CDC, 2022 (*12*) |
|  |  | SCOV2-Rev | CCTTGTGTGGTCTGCATGAGTTTAG |  |
|  |  | SCOV2-Probe | (FAM)-ATTGCAACAATCCATGAGCAGTGCTGACTC-(BHQ) |  |
| *Staphylococcus aureus* | A | SA-Fwd | GTTGCTTAGTGTTAA[+C]TTTAGTTGTA | Chochua *et al*, 2016 (*13*) |
|  |  | SA-Rev | AATGTCGCAGGTTCTTTATGTAATTT |  |
|  |  | SA-Probe | (FAM)-AAGTCTAAGTAGCTC[+A]GCAAATGCA-(BHQ) |  |
| Spn (*lytA*) | A | *lytA*-Fwd | ACGCAATCTAGCAGATGAAGCA | Carvalho *et al*, 2007 (*14*) |
|  |  | *lytA*-Rev | TCGTGCGTTTTAATTCCAGCT |  |
|  |  | *lytA*-Probe | (FAM)-GCCGAAAACGCTTGATACAGGGAG-(BHQ) |  |
| Spn (*piaB*) | A | *piaB*-Fwd | CATTGGTGGCTTAGTAAGTGCAA | Trzcinski et al, 2013 (*15*) |
|  |  | *piaB*-Rev | TACTAACACAAGTTCCTGATAAGGCAAGT |  |
|  |  | *piaB*-Probe | (FAM)-TGTAAGCGGAAAAGCAGGCCTTACCC-(BHQ) |  |
| Spn ST 3 | B | 3-Fwd | GGTCAGCAGAAAGTATGCATTGG | Azzari et al, 2010 (*16*) |
|  |  | 3-Rev | TCGTTTATCCAGGGTCTGATGA |  |
|  |  | 3-Probe | (FAM)-TATTGGATGTGGTTTATCG[+T]GAAGA-(BHQ) |  |
| Spn SG 6A/B/C/D | A | 6A/B/C/D-Fwd | AAGTTTGCACTAGAGTATGGGAAGGT | Azzari et al, 2010 (*16*) |
|  |  | 6A/B/C/D-Rev | ACATTATGTCCRTGTCTTCGATACAAG |  |
|  |  | 6A/B/C/D-Probe | (FAM)-TGTTCTGCCCTGAGCAACTGG-(BHQ) |  |
| Spn SG 6C/D | B | 6C/D-Fwd | TTGGGATGATTGGT[+C]GTATTAG | Pimienta et al, 2013 (*17*) |
|  |  | 6C/D-Rev | CTCTTCAATTAGTT[+C]TTCAGTTCG |  |
|  |  | 6C/D-Probe | (FAM)-CCACGCAATTCGCCATC-(MGB) |  |
| Spn SG 7A/F | B | 7A/F-Fwd | GATGGCATGTGGCAAACCA | Azzari et al, 2010 (*16*) |
|  |  | 7A/F-Rev | TTTGCCCTCCTTAATCATTTCAC |  |
|  |  | 7A/F-Probe | (FAM)-TTGGCTATCGGCATGGTGGT-(BHQ) |  |
| Spn ST 7C | A | 7C-Fwd | CGTCAGGAATAGGTGCAATCTCT | Olwagen *et al*, 2017 (*18*) |
|  |  | 7C-Rev | TGAAATTCCAAGCGAAGCAA |  |
|  |  | 7C-Probe | (FAM)-TTCATCTATTGGTTCTT[+A]TGGT[+G]TT-(BHQ) |  |
| Spn ST 8 | A | 8-Fwd | CCACTCATCAGTTTCCCATATGTTT | Azzari et al, 2010 (*16*) |
|  |  | 8-Rev | TCAATAATTGAAGAAGCGAACGTT |  |
|  |  | 8-Probe | (FAM)-TGATGGCAGATGGGTTGGGACGAG-(BHQ) |  |
| Spn SG 9A/L/N/V | A | 9A/L/N/V-Fwd | TGGAATGGGCAAAGGGTAGTA | Azzari et al, 2010 (*16*) |
|  |  | 9A/L/N/V-Rev | TCGGTTCCCCAAGATTTTCTC |  |
|  |  | 9A/L/N/V-Probe | (FAM)-TTAATCATGCTAACGGCTCA[+T]CGA-(BHQ) |  |
| Spn ST 10A | B | 10A-Fwd | CCTCTCCTATCAACTATTACTCATTATACTACCT | Azzari et al, 2010 (*16*) |
|  |  | 10A-Rev | AATAACCATAAGTCCCTAGATCATTCAAAG |  |
|  |  | 10A-Probe | (FAM)-TCATTACAACTCCCTATGTGACACGGGTCTTTT-(BHQ) |  |
| Spn SG 11A/B/C/D/F | B | 11A/B/C/D/F-Fwd | ACCGCATTTCTTATCGCACTATATT | Olwagen *et al*, 2017 (*18*) |
|  |  | 11A/B/C/D/F-Rev | TCTCCTTACCATCAAACATGTTAATCA |  |
|  |  | 11A/B/C/D/F-Probe | (FAM)-TGAATCAGTCTGACCGTTT-(MGB) |  |
| Spn SG 12A/12B/12F/44/46 | B | 12A/12B/12F/44/46-Fwd | GATTATTCGCTTGCCTCTTCATG | Azzari et al, 2010 (*16*) |
|  |  | 12A/12B/12F/44/46-Rev | ATAGCCGAAATAAGCTTTCCAGAA |  |
|  |  | 12A/12B/12F/44/46-Probe | (FAM)-ATTTGTAAGCGGACGTGCGATT-(BHQ) |  |
| Spn SG 15A/B/C/F | B | 15A/B/C/F-Fwd | TTGAATCAGGTAGATTGATTTCTGCTA | Azzari et al, 2010 (*16*) |
|  |  | 15A/B/C/F-Rev | CTCTAGGAATCAAATACTGAGTCCTAATGA |  |
|  |  | 15A/B/C/F-Probe | (FAM)-CTCCGGCTTTTGTCTTCT[+C]TGT-(BHQ) |  |
| Spn SG 16F | B | 16F-Fwd | GCAACTGGTATTTTTGATATTGGAGAA | Olwagen *et al*, 2017 (*18*) |
|  |  | 16F-Rev | CAAAGGAATGCCATGCCATA |  |
|  |  | 16F-Probe | (FAM)-AAAATGCTAACTTCGTTGGA[+G]G-(BHQ) |  |
| Spn SG 18A/B/C | B | 18A/B/C-Fwd | CCTGTTGTTATTCACGCCTTACG | Azzari et al, 2010 (*16*) |
|  |  | 18A/B/C-Rev | TTGCACTTCTCGAATAGCCTTACTC |  |
|  |  | 18A/B/C-Probe | (FAM)-AACCGTTGGCCCTTGTGGTGGA-(BHQ) |  |
| Spn ST 19A | A | 19A-Fwd | TTCGACGACGTATCAGCTTCA | Azzari et al, 2010 (*16*) |
|  |  | 19A-Rev | TCATTGAGAGCCTTAACCTCTTCA |  |
|  |  | 19A-Probe | (FAM)-ACCCAAAACGGTTGACGCATTATACT-(BHQ) |  |
| Spn SG 19B/F | A | 19B/F-Fwd | GGTCATGCGAGATACGACAGAA | Azzari et al, 2010 (*16*) |
|  |  | 19B/F-Rev | TCCTCATCAGTCCCAACCAATT |  |
|  |  | 19B/F-Probe | (FAM)-ACCTGAAGGAGTAGCTGCTGGAACGTTG-(BHQ) |  |
| Spn ST 20 | B | 20-Fwd | AAAGATACTGGCTGAGGAGCTATCTATT | Azzari et al, 2010 (*16*) |
|  |  | 20-Rev | AGTCAAAAGTACTCAACCATTCTGATATATTC |  |
|  |  | 20-Probe | (FAM)-AGGATAAGGTCTA[+C]TTTGTGGGAGTTC-(BHQ) |  |
| Spn ST 21 | A | 21-Fwd | CCATTTGAAGGACCAGTTGTTG | Olwagen *et al*, 2017 (*18*) |
|  |  | 21-Rev | AAAAAGCCACTATCAGGAATACCAA |  |
|  |  | 21-Probe | (FAM)-AATGGCATTGCTTCGTAAA-(MGB) |  |
| Spn ST 22A/F | B | 22A/F-Fwd | TCTATTAAATAACCCATTGGAATTGAAACG | Miellet et al, 2022 (*19*) |
|  |  | 22A/F-Rev | TCGCAATTGAAGACCACATAAACTG |  |
|  |  | 22A/F-Probe | (FAM)-TCCGTAATGCGCTTATGAGCACATTCTCCA-(BHQ) |  |
| Spn SG 23A/B/F | A | 23A/B/F-Fwd | GGTGGACTTTCCGATGCAA | Olwagen *et al*, 2017 (*18*) |
|  |  | 23A/B/F-Rev | CACTGTCAACAAAAATGAGGTAATCTC |  |
|  |  | 23A/B/F-Probe | (FAM)-AAATGTCGGTATAGATAAAG-(MGB) |  |
| Spn SG 25A/25F/38 | A | 25A/25F/38-Fwd | GTCTTACGTAGAACCTCTCTGGATGA | Azzari et al, 2010 (*16*) |
|  |  | 25A/25F/38-Rev | TGGTCCTACAAGCGACATGTG |  |
|  |  | 25A/25F/38-Probe | (FAM)-TTGCCACAGATTTGGAATATTTTGGTCGG-(BHQ) |  |
| Spn SG 33A/F/37 | B | 33A/F/37-Fwd | CGAGAGAGAATATGAGGGAATTGTTA | Azzari et al, 2010 (*16*) |
|  |  | 33A/F/37-Rev | TCTCAATCCCCGCATTTACTG |  |
|  |  | 33A/F/37-Probe | (FAM)-AGGAAAACTGTGGTCACGGTTCG-(BHQ) |  |
| Spn SG 34/37/17A | A | 34/37/17A-Fwd | GGATACTATGTACGAACAGATGGACTTG | Olwagen *et al*, 2017 (*18*) |
|  |  | 34/37/17A-Rev | CTCACTAACTCGCCCGAATAAAC |  |
|  |  | 34/37/17A-Probe | (FAM)-CCGACTATACTCCATTTGA-(MGB) |  |
| Spn ST 35B | B | 35B-Fwd | GCATGGAGGTGGAGCATACA | Azzari et al, 2010 (*16*) |
|  |  | 35B-Rev | TGTAAAGACTGCACAACTCGATATAAAA |  |
|  |  | 35B-Probe | (FAM)-CAATTTAAACAATATTAGTAAAGCGCAGGTCAAGCAAA-(BHQ) |  |
| *Streptococcus pyogenes* | B | SPY-Fwd | GCACTCGCTACTATTTCTTACCTCAA | CDC 2008 and |
|  |  | SPY-Rev | GTCACAATGTCTTGGAAACCAGTAAT | Olwagen *et al*, 2017 (*18*) |
|  |  | SPY-Probe | (FAM)-CCGCAACTCATCAAGGATTTCTGTTACCA-(BHQ) |  |
| Respiratory syncytial virus A | C | RSVA-Fwd | AGATCAACTTCTGTCATCCAGCAA | van der Pol *et al*, 2010 (*20*) |
|  |  | RSVA-Rev | TTCTGCACATCATAATTAGGAGTATCAAT |  |
|  |  | RSVA-Probe | (FAM)-CACCATCCAACGGAGCACAGGAGAT-(BHQ) |  |
| Respiratory syncytial virus B | C | RSVB-Fwd | AAGATGCAAATCATAAATTCACAGGA | van der Pol *et al*, 2010 (*20*) |
|  |  | RSVB-Rev | TGATATCCAGCATCTTTAAGTATCTTTATAGTG |  |
|  |  | RSVB-Probe | (FAM)-TTCCCTTCCTAACCTGGACATAGCATATAACATACCT-(BHQ) |  |

**Table S3.** gBlock (external synthetic calibrator) sequences (5’-3’). Spn = *Streptococcus pneumoniae.*

A1

| Targets | Sequence |
| --- | --- |
| *Hemophilus influenzae*  *Moraxella catarrhalis*  *Staphylococcus aureus*  Spn *(lytA)*  Spn *(piaB)*  Spn *19B/F*  Spn *23A/B/F*  Spn *25A/F/38*  Spn *34/37/17A* | TCA CAT TGG CTA ATC ACT TGG GCA AAA TTG CCA AGA TTA AAT GCT TCT TAG CCT GCG GTT AAA CCG AGA CTT CTT TGT GCA GTA TGG CGA GCA TTA TTC CGG CCA ATT AAT GAC GCG GCG TGT TGA CCG TTT TGA CTT TCG AAG TTC ACC GAC ATC AAC CCA AGC TTT GGT AAT TGC CGC GTC AGC GGT AAC CTA ATC TAA GCA GCC GTA GTT TTG TCG CTC AAA GTT GCT TAG TGT TAA CTT TAG TTG TAG TGT ACA TCA AGT CTA AGT AGC TCA GCA AAT GCA TTT AGT GCC AAA ATT ACA TAA AGA ACC TGC GAC ATT TTT TTC GTC CGC AAT AAG CGC AGA GAC GCA ATC TAG CAG ATG AAG CAT TCG GAA CGC CGA AAA CGC TTG ATA CAG GGA GTA CTG AGC TGG AAT TAA AAC GCA CGA CTA GAT CAA CAG TTG TTG GCC GAC TCA TTG GTG GCT TAG TAA GTG CAA ATT GGC ATC TGT AAG CGG AAA AGC AGG CCT TAC CCT AAT GGC CAC TTG CCT TAT CAG GAA CTT GTG TTA GTA CTT CGG TCA AGG AAC TCT GAG TAT CGG TCA TGC GAG ATA CGA CAG AAT GCC TTT AGA ACC TGA AGG AGT AGC TGC TGG AAC GTT GCG ATT TAA TTG GTT GGG ACT GAT GAG GAG AAG AGA CGT TCG CTA GCC TAT CTT GGT GGA CTT TCC GAT GCA ATC TAT TGA GCA AAT GTC GGT ATA GAT AAA GTG TCA TTG AGA TTA CCT CAT TTT TGT TGA CAG TGT TTC GTA CGC AGG TTA ACG ACG CTA GTC TTA CGT AGA ACC TCT CTG GAT GAT TAG GAT CCT TTG CCA CAG ATT TGG AAT ATT TTG GTC GGT TTC TGA CAC ATG TCG CTT GTA GGA CCA CAT ACT AGT AGC ATG GCT ACT CGT GGG ATA CTA TGT ACG AAC AGA TGG ACT TGA TCA GTG CCC GAC TAT ACT CCA TTT GAA TCT TCG AGT GTT TAT TCG GGC GAG TTA GTG AGA CCG CGT CTC TAG CGT GGG AGA TAA TCC TTA TTA T |

A2

| Targets | Sequence |
| --- | --- |
| Spn 6A/B/C/D  Spn 9A/L/N/V  Spn 19A  Spn 21  Spn 7C  Spn 8  *Mycoplasma pneumoniae*  Adenovirus | CTG GAA TCT ATG ATA TGC GCT AGT TCA TGA CGG GAC ACC CTA AGT TTG CAC TAG AGT ATG GGA AGG TTG CTA TTG TTC TGC CCT GAG CAA CTG GTT TCC TGA GAC TTG TAT CGA AGA CAC GGA CAT AAT GTT AAA GTC TCA GTC GAG TTG TCA GCC TGG AAT GGG CAA AGG GTA GTA TTT CGA TTT AAT CAT GCT AAC GGC TCA TCG ATC TAG GAG AAA ATC TTG GGG AAC CGA CGC AAG CTG TGA TAT ATG TCA CTG CTT CGA CGA CGT ATC AGC TTC AAT GCG ACT TAC CCA AAA CGG TTG ACG CAT TAT ACT GTA TTC TGA AGA GGT TAA GGC TCT CAA TGA AAC GCC GCG TGA GAT TTA TAC TCT GCC ATT TGA AGG ACC AGT TGT TGC TTG ACT TGA AAT GGC ATT GCT TCG TAA AGT AAC TCG TTG GTA TTC CTG ATA GTG GCT TTT TGA AGG ATC AAT ACT GCC GTC CTG TTC GTC AGG AAT AGG TGC AAT CTC TGT TTC ATT TCA TCT ATT GGT TCT TAT GGT GTT ACT TGG TAC TTG CTT CGC TTG GAA TTT CAC TGA CGA CTC AAG CAG CTG ATT TGT CCA CTC ATC AGT TTC CCA TAT GTT TAC GTT TAG TCT GAT GGC AGA TGG GTT GGG ACG AGC TAT TCG GAA ACG TTC GCT TCT TCA ATT ATT GAT TTC CGG TAT CAG CAG CGT AAA GTC GGT CAA TCT GGC GTG GAT CTT ATT TGC TCC CCC GTT GAA AAA GTG AGT GGG TAG TTT CGC TTG TGG GGC AGT TAC CAG TCG CTT GCA CGG TTA CTC TGA AAA TTT GAG GTG GAT CCA ATG GAC ATG CTT GTA ACC ACG TCG AAA ACT TCG AAT TAT CGT ACG TAT CTG CGT ACA CCG TTC TAG CGT ACG TGA GTC CCT ACT TAG ATG CTT A |

B1

| Targets | Sequence |
| --- | --- |
| 16S  Spn 11A/B/C/D/F  Spn 12A/12B/12F/44/46  Spn 15A/B/C/F  Spn 16F  Spn 18A/B/C  Spn 6C/D  Spn 7A/F  Spn 10A | GAG CTT CAA GGA GCA TTC CGT GTC AAG CTA TCT CCT ACG GGA GGC AGC AGT TAG TCC GTA TTA CCG CGG CTG CTG GCA CAC GAG TCT TAA CAG GAT TAG ATA CCC TGG TAG TCC AGA TGC TTC GTG CAT CGC ATA TAC GAC CGC ATT TCT TAT CGC ACT ATA TTA GTC TTT GAA TCA GTC TGA CCG TTT GCT TAT GAT TAA CAT GTT TGA TGG TAA GGA GAC CTG GGC TTA CAT ACT TGA GAG ATC GAT TAT TCG CTT GCC TCT TCA TGG TCT TTA ATT TGT AAG CGG ACG TGC GAT TCT TGA CTT GAT TCT GGA AAG CTT ATT TCG GCT ATA CCC GCG TTG ATG TTA CTG TCA AGA TTG AAT CAG GTA GAT TGA TTT CTG CTA TTT CGG ACT ACT CCG GCT TTT GTC TTC TCT GTT TTG TCA TCA TTA GGA CTC AGT ATT TGA TTC CTA GAG CAT ACT ACG TGG ATA TCC GGG ATC TGC AAC TGG TAT TTT TGA TAT TGG AGA ATA GTC AAA ATG CTA ACT TCG TTG GAG GGT TCA TTT ATG GCA TGG CAT TCC TTT GTT ATG GCA CTA CCG ATG GTG ACA TCC CTG TTG TTA TTC ACG CCT TAC GAC TGT AAC CGT TGG CCC TTG TGG TGG AAT CTG GAG TAA GGC TAT TCG AGA AGT GCA ATA CTG CAA CCA GGA TGT CGA CTT GTT TGG GAT GAT TGG TCG TAT TAG AGC TTA CGT CCA CGC AAT TCG CCA TCA GTG CTA CCG AAC TGA AGA ACT AAT TGA AGA GTT ACA GGC TAA TCA CTA GCG CTG GTG ATG GCA TGT GGC AAA CCA TTT AGC TTT GGC TAT CGG CAT GGT GGT TAT GCA GCG TGA AAT GAT TAA GGA GGG CAA AAT AGT GCG TGT TCA TCA ACT CCG AGC CTC TCC TAT CAA CTA TTA CTC ATT ATA CTA CCT CTA GTA TTC GTC ATT ACA ACT CCC TAT GTG ACA CGG GTC TTT TTT TCG TAC TTT GAA TGA TCT AGG GAC TTA TGG TTA TTG TTC AGA GCA TCG AGC GCT ATT TAT AGC AGC C |

B2

| Targets | Sequence |
| --- | --- |
| Spn 13  Spn 20  Spn 22A/F  Spn 3  Spn 35B  Spn 33A/F/37  *Streptococcus pyogenes*  Bocavirus | AGC TTC CCG AAT GAC ACG ATA AAG GTC GCC ATT GCG TTG TTT CGG ATT TAG TAG TAA CCC CAT TGA TCT ACA GTT GAG TAG TAA GAG ATC ATA TTC AAG CTT GAC GAT GGA AAT GCA TCC TCA ATC AAG AAA CGT CAT AGG AGT CGG ATC TTT CCA AAA GAT ACT GGC TGA GGA GCT ATC TAT TTG CAA CGT AGG ATA AGG TCT ACT TTG TGG GAG TTC TGG CAC ATG AAT ATA TCA GAA TGG TTG AGT ACT TTT GAC TCT GGG TCT ATA GCG TAT CAC GCA TAT CTA TTA AAT AAC CCA TTG GAA TTG AAA CGT CAT TGT TCC GTA ATG CGC TTA TGA GCA CAT TCT CCA CTG TAC AGT TTA TGT GGT CTT CAA TTG CGA TTT AAT GCA GTA CGT CGG CCG AAT CGG TCA GCA GAA AGT ATG CAT TGG ATT CGT TTA TTG GAT GTG GTT TAT CGT GAA GAA TTC GTT TCA TCA GAC CCT GGA TAA ACG ATG CGT TCA GAT TAT TGC CAG CCG AAG CAT GGA GGT GGA GCA TAC AGT TAC TCA ATT TAA ACA ATA TTA GTA AAG CGC AGG TCA AGC AAA TTG GCT CAA TTT TTA TAT CGA GTT GTG CAG TCT TTA CAG ATT TAA GCA TCG TCA GCT GGC ATC CGA GAG AGA ATA TGA GGG AAT TGT TAC CTG ATG AAG GAA AAC TGT GGT CAC GGT TCG TCT TAC TGA GCA GTA AAT GCG GGG ATT GAG AAA TCG TAA GTG CTT GTC GGC CAA TCG CAC TCG CTA CTA TTT CTT ACC TCA AAG GTC CAT TCC GCA ACT CAT CAA GGA TTT CTG TTA CCA TGC ATT CAG ATT ACT GGT TTC CAA GAC ATT GTG ACG ATT CGA CTG CTC ATA CCT GAG GAT GGA AGA GAC ACT GGC AGA CAA TAT GTT CCT GCG GCT CCT GCT CCT GTG ATG TTC TTA GCT CAT ATC ATC AGG AAC ACC CAC CGA TTT AAC CAG TTC GGG ACA GGC TTT |

C1

| Targets | Sequence |
| --- | --- |
| SARS-CoV-2  Influenza A  Influenza B  RSV A  RSV B  Human Rhinovirus  EAV | GCC GAA CTG TAC ATG TAC TCG TAT GTC GAT ACT GCA GAT TTG GAT GAT TTC TCC TCT GCG AAA TTG CAA CAA TCC ATG AGC AGT GCT GAC TCA TGT CCT AAA CTC ATG CAG ACC ACA CAA GGC TCT ATG CTA CTG AGG CAC GTA GTA AAG ACC AAT CCT GTC ACC TCT GAT CGT ATT TGT GTT CAC GCT CAC CGT GCC TTC AGT GGA CTG CAG CGT AGA CGC TTT GCT AAG TTC CCA CTT TGG AAG CTG AGA AAT ACG GTG GAT TAA ACA AAA GCA AAA TTC CGG CAC CCA TAT TGG GCA ATT TCC TAT GGC GGT TAA ATT TCT TCG GAG CTA TTG CTG GCT GAG GAT GCT CAT ACG AAT AGG ACA GAT CAA CTT CTG TCA TCC AGC AAT TTG AGT CCA CAC CAT CCA ACG GAG CAC AGG AGA TAC TAT CGG CAC TAT AAA GAT ACT TAA AGA TGC TGG ATA TCA CAC TTG GGA GCT CCA CAT AAT CCA ACT AGC CTG CGT GGC GTA CAA TCC TCC GGC CCC TGA ATG TGG CTG TCC GAA ATA CTT TGG GTG TCC GTG TTT CCA CAA CTT TCG AGC ATT TAA GCC CGG GCG ACA GCC TAC AAG CTA CAC ATA CTC TGA TTG CGG ACC CGC ATC TGA CCA ATG GAA CTC TTC ACT CAC TGC AGA TGC CGC TTT GAG CGT TGT AAG ACC CA |

C2

| Targets | Sequence |
| --- | --- |
| Coronavirus 229E  Coronavirus NL63  Coronavirus OC43  Coronavirus HKU  Parainfluenzavirus 1  Enterovirus  Parainfluenzavirus 3  Parainfluenzavirus 2  Parainfluenzavirus 4 | ACA TAC CGT GAC CCT GGA CCG ACT TTG TCA GTT CGT ATG CTA AAC GAG CAT TCA TTA TCA GCT TAT GAC TTG GCG TGT TAG ACG TGT TAA TGC CAA TGT TCT GAC AAT CGC GCG TTG ATG TTA CTG TCA AGA GCG TGT TCC TAC CAG AGA GGA CGT ATA TGT TAT TCA GTG CTT TGG TCC TCG TGA TAT CCT GAC TGC CAA AGG TTT TCC ACA GCC CGA TTG CTG GAG TTA CAT AAC CGT CGA TGA GGC TAT TCC GAC TAG GTA GCT TAC GTT CCG CCT GGC ACG GTA CTC CCT GTC TAG GTT ACT ATA TTG AAG GCT CAG GAA GGA TAC TGA GCC GTC CTC TGA ACA CTT AGT TCC CAT TGC TTT CGG AGT AAG GAC TCT CCC CTT CTG AAG CAA TCG TCG TAT TGG ATA TTG GTA TAG ACA CAG CCG GGG CGC GGA GAT CAC AAC TTT TTA TCT GAT TTA AAC CCG GTA ATT TCT CAT CGT ACG TAA CGA CAA CAG GAA ATC TCT GCA GAT TCT GTA ATA GCT GCA GGA ACA AGG TGG TCT GAC TGC GTG TTG ATA TCT CCC TGA ATG CGG CTA ATC CAT AGC ACC AAC CGA CTA CTT TGG GTG TCC GTG TTT CGA TAT TCC GTG GCT GCT TAT GGT GAC AAT TCA CCA ATG TGC AGC GGT TAC TGA TTG ATG AAA GAT CAG ATT ATG CAT GAT CAG CTT GGA CCA GGG ATA TAC TAC AAA GGC AAA ATA ATA TTT CTC CAT TGT TCA CAA CTG GGT GTC CCG GTG AAT TCT GCG AGT GCA CAA TGC CTA GGA CTA TGA AAA CCA TTT ACC TAA GTG ACG TTA TAT CAA TCG CAA AAG CTG TTC AGT CAC TGC TAT ACA TGA GCC TTG ATC TAG CTG AAC TGA GAC TTG CTT ACC CGC GTT GAT GTT ACT GTC AAG ACA AAC GAT CCA CAG CAA AGA TTC CTT GAC TTG AGT ATC ATC ATC TGC CAA ATC GGC AAT TAA ACA TTA TGC TGC TTT CCT TAC AGG CCA CAT CGT ATT CGA TAT AGG TGA CCT ATG AAA GCT GGC CCT CT |
